# α2,6-Sialylation is Upregulated in Severe COVID-19 Implicating the Complement Cascade

**DOI:** 10.1101/2022.06.06.22275981

**Authors:** Rui Qin, Emma Kurz, Shuhui Chen, Briana Zeck, Luis Chiribogas, Dana Jackson, Alex Herchen, Tyson Attia, Michael Carlock, Amy Rapkiewicz, Dafna Bar-Sagi, Bruce Ritchie, Ted M. Ross, Lara K. Mahal

## Abstract

Better understanding of the mechanisms of COVID-19 severity is desperately needed in current times. Although hyper-inflammation drives severe COVID-19, precise mechanisms triggering this cascade and what role glycosylation might play therein is unknown. Here we report the first high-throughput glycomic analysis of COVID-19 plasma samples and autopsy tissues. We find α2,6-sialylation is upregulated in plasma of patients with severe COVID-19 and in the lung. This glycan motif is enriched on members of the complement cascade, which show higher levels of sialylation in severe COVID-19. In the lung tissue, we observe increased complement deposition, associated with elevated α2,6-sialylation levels, corresponding to elevated markers of poor prognosis (IL-6) and fibrotic response. We also observe upregulation of the α2,6-sialylation enzyme ST6GAL1 in patients who succumbed to COVID-19. Our work identifies a heretofore undescribed relationship between sialylation and complement in severe COVID-19, potentially informing future therapeutic development.

## INTRODUCTION

COVID-19, the clinical syndrome caused by SARS-CoV-2 infection, varies in severity from mild respiratory symptoms, to pneumonia requiring hospitalization, to death^1^. Over the last two years, the immune response in COVID-19 has been studied in effort to characterize disease pathology and better understand the potential therapeutic options for severe COVID-19^2^. One of the milestones of severe COVID-19 is hyper-inflammation, which are associated with acute respiratory distress syndrome (ARDS), dysregulation of cytokine release, NFκB signaling and immune cell mobilization, thrombosis, increased vascular permeability, and endothelial damage^3–7^. The complement cascade is an important trigger of inflammation and has been shown activated in COVID-19 in multiple studies^8^. This proteolytic cascade produces various pro-inflammatory molecules and results in the formation of the membrane attack complex (MAC) that causes cell death and tissue damage^9^. In severe COVID-19, the augmented inflammatory signatures, including formation of neutrophil extracellular traps (NETs), increased myeloid cell recruitment and higher cytokine levels, all map onto dysregulation of the complement cascade^10^. Therefore, there has been an increasing interest in evaluating the therapeutics for severe COVID-19 that specifically target the complement cascade^11–13^.

Glycosylation plays essential and increasingly appreciated roles in regulating inflammation and immune response in infectious diseases^14–17^. For example, one of the best studied roles for glycans in inflammation is the recruitment of leukocytes to sites of injury via recognition by selectins of glycans such as sialyl Lewis x^18, 19^. Antibody-mediated immune response is also impacted by glycosylation. In IgG, core fucosylation inhibits interactions with Fc receptors, diminishing antibody-dependent cellular cytotoxicity (ADCC)^15, 20, 21^. Several other glycan features, including sialylation, bisecting GlcNAc, and galactosylation, have also been associated with IgG function^15, 22^. Glycosylation also plays a key role in the host immune response to pathogens, which determines the disease severity^14^. In recent work on influenza, severe disease was found to be associated with high expression of high mannose in the lung, a glycoform that binds the innate immune receptor mannose binding lectin (MBL2)^23^. In COVID-19, glycosylation patterns of SARS-CoV-2-specific antibodies correlated with severity. Lower core fucosylation was associated with higher severity, in line with the impact of this modification on ADCC^24, 25^. Although IgG glycosylation was studied in COVID-19, there has been no work examining glycosylation in whole plasma or in affected tissues.

In this work, we utilized high-throughput lectin microarray technology to examine glycosylation as a function of COVID-19 severity in both plasma and autopsy tissues, with a focus on identifying glycomic markers of severity and understanding their potential roles. We found higher levels of α2,6-sialylation in the plasma of severe COVID-19 patients, which was also observed in the lower lobes of the lungs in patients who succumbed to COVID-19. In the severe cases, plasma glycoproteins bearing this epitope were enriched in members of the complement cascade, which had a greater fraction of α2,6-sialylated protein compared to the mild cases. Complement deposition and increased myeloid recruitment were observed in the lower lobe lungs of COVID-19 autopsies. We also identified higher levels of ST6GAL1, the main enzyme that biosynthesizes α2,6-sialic acid, in COVID-19 patients. Overall, our work points to a previously unexplored role of α2,6-sialylation in complement system biology. This newly discovered association may have important consequences for the development of therapeutic approaches to ameliorate the detrimental immune responses resulting from the overactivated complement cascade in severe COVID-19.

## METHODS

### Cohorts and Sample Collection

COVID-19 plasma samples were collected from 71 patients recruited from the Intensive Care Unit, the hospital ward or the outpatient clinic at the University Hospital (Edmonton, Alberta, Canada). The CoCollab study was reviewed and approved by the Research Ethics Board/Alberta Research Information Servies (ARISE) at the University of Alberta. Recruits were informed of the details of the study by the study team, had the opportunity to ask questions, then signed informed consent. Plasma samples analyzed in this study were collected at the time of enrollment. Blood samples were processed within one hour where possible to isolate plasma, and peripheral blood mononuclear cells, then aliquoted into 100 microliter cryovials.

Non-COVID-19 plasma samples were collected from 60 adults originally recruited for a study of influenza vaccination response among the general population, at the University of Georgia Clinical and Translational Research Unit (Athens, Georgia, USA) from September 2019 to December 2019. All volunteers were enrolled with written, informed consent. Participants were excluded if they, at the time of enrollment, already received the seasonal influenza vaccine, had acute or chronic conditions that would put the participant at risk for an adverse reaction to the blood draw or the flu vaccine (e.g., Guillain-Barré syndrome or allergies to egg products), or had conditions that could skew the analysis (e.g., recent flu symptoms or steroid injections/medications). Plasma samples analyzed in this present study were collected prior to vaccination. A brief description of the two cohorts above is in **Supplementary Table 1**.

Hospital-based autopsies for COVID-19 patients were performed at NYU Winthrop Hospital (Mineola, New York, USA) among persons with laboratory-confirmed COVID-19 or who were under investigation and tested positive on post-mortem PCR. Autopsies were performed between the dates of March 2020 and April 2020. The lungs, heart, kidneys, and liver were used in this study. Tissues were fixed in 10% buffered formalin for 72 h and routinely processed. Details about the clinical characteristics of the COVID-19 cohort and of matched COVID-19 negative cohort can be found in **Supplementary Table 2**.

### Fluorescent Labelling of Samples

Total protein concentrations of plasma and autopsy samples were determined with *DC*™ protein assay (Bio-Rad Laboratories). PBS refers to phosphate-buffered saline (137 mM NaCl, 2.7 mM KCl, 8.9 mM Na_2_HPO_4_, and 1.8 mM KH_2_PO_4_, pH = 7.4) hereinafter. PBST refers to PBS supplemented with Tween® 20 (concentration in v/v indicated where it appears) hereinafter.

To label plasma proteins, each sample containing 10 μg total protein was first diluted in PBS to 27 μl. The pH of the solution was adjusted with 3 μL of 1M sodium bicarbonate. Then 0.21 μl of a 10 mg/ml Alexa Fluor™ 555 NHS ester (Thermo Fisher Scientific) stock solution was thoroughly mixed with the sample solution. The mixture was incubated in the dark and at room temperature with gentle agitation. After 1 hour, unconjugated dyes were removed by Zeba™ dye and biotin removal filter plates (Thermo Fisher Scientific). The reference standard, a commercial human plasma (Millipore Sigma, catalog #P9523), was fluorescently labelled with Alexa Fluor™ 647 NHS ester (Thermo Fisher Scientific) in a similar fashion. The amounts of reagents were scaled linearly to the starting protein amount (2 mg). Finally, each Alexa Fluor™ 555-labeled sample (10 μg of total protein) was mixed with a proper volume of Alexa Fluor™ 647-labeled reference standard containing the same amount of protein. The dual-color mixture was first diluted to 50 μl with PBS then mixed with 50 μl 0.1% PBST.

To label tissue samples from autopsy, each sample containing 50 μg total protein was first diluted in PBS to 60 μl. The pH of the solution was adjusted with 6.7 μL of 1M sodium bicarbonate. Then 0.2 μl of a 10 mg/ml Alexa Fluor™ 555 NHS ester (Thermo Fisher Scientific) stock solution was thoroughly mixed with the sample solution. The mixture was incubated in the dark and at room temperature with gentle agitation. After 1 hour, unconjugated dyes were removed by Zeba™ dye and biotin removal filter plates (Thermo Fisher Scientific). The pool reference was generated and fluorescently labelled with Alexa Fluor™ 647 NHS ester (Thermo Fisher Scientific) in a similar fashion. Then, each Alexa Fluor™ 555-labeled sample (3 μg of protein) was mixed with a proper volume of Alexa Fluor™ 647-labeled reference standard containing the same amount of protein. The dual-color mixture was first diluted to 74 μl with PBS then mixed with 2 μl 0.2% PBST.

### Fabrication of Lectin Microarray Slides

Lectin microarray slides were fabricated as previously described^26^ in a published protocol. In brief, lectins and antibodies were printed on Nexterion^®^ Slide H (Applied Microarrays) with the microarray printer Nano-Plotter™ 2.1 (GeSim). The temperature and humidity inside the printer chamber were maintained at 14℃ and 50%, respectively. Inhibiting sugars were added to lectin solutions to a final concentration of 50mM (except lactose: 25mM) prior to printing. Lectins for printing, concentrations and inhibiting sugars are listed in **Supplementary Table 3** (for plasma samples) and **Supplementary Table 4** (for autopsy samples).

### Dual-color Lectin Microarray

All steps were performed in the dark at room temperature. Each dual-color mixture was allowed to hybridize with the microarrays for 1 hour. Microarrays were washed twice with 0.005% PBST for 10 minutes and once with PBS for 5 minutes. The slides were briefly rinsed with ultrapure water and dried by centrifugation. Fluorescence signals were gained with Genepix™ 4400A fluorescence slide scanner (Molecular Devices) in the 532 nm channel and the 635 nm channel that correspond to the excitation/emission profiles of Alexa Fluor™ 555 and Alexa Fluor™ 647, respectively. Raw fluorescence signals and background signals were generated by the Genepix Pro™ 7 software (Molecular Devices), which were further processed and analyzed with a custom script as previously described^27^. Heatmaps, boxplots and volcano plots were generated with R (Version: 4.0.1).

### Lectin Pulldown of Plasma Samples

In this section, centrifugation (1000x *g*, 2 minutes) was used to remove liquid from columns in washes and elutions. All steps were performed at room temperature. To prepare SNA-agarose columns, 200 μl 50% suspension of streptavidin-agarose resin (Millipore Sigma), was added to each microcentrifuge column. Storage buffer was removed, and the resin was washed with 200 μl PBS. 400 μl of biotinylated *Sambucus nigra* lectin (SNA, Vector Laboratories, pre-diluted to 0.5 mg/ml with PBS) was added to the column and the mixture was incubated with gentle agitation for 30 minutes. Then the resin was washed with 200 μl PBS twice. Control columns were prepared using the same procedure except that 400 μl of PBS was added to the column instead of biotinylated SNA.

To prepare SNA pulldown samples for mass spectrometry analysis, pooled plasma samples corresponding to the mild and severe COVID-19 patient group were prepared by combining equal volumes of individual samples. Each pooled plasma sample containing 300 μg of total protein was diluted to 300 μl with PBS. Pulldown was performed in triplicates (i.e., each pooled sample was enriched with three separate columns prepared with the same procedure at the same time). Diluted samples were incubated with the SNA-bound resin or the control resin for 1 hour with gentle agitation. The resin was washed with 400 μl PBS three times. To elute glycoproteins, 75 μl of 0.2 M lactose in PBS was added to the column and incubated with gentle agitation. After 30 minutes, the flow-through was collected. Then 75 μl of 0.2 M lactose in 0.2 M acetic acid was added to the column and incubated with gentle agitation. After 30 minutes, the flow-through was collected and combined with the previous flow-through. Finally, the pH of the combined eluate was adjusted with 1M Tris (pH = 9.0) to 7.5.

To prepare SNA pulldown samples for western blotting, pooled plasma samples corresponding to the mild COVID-19, severe COVID-19 and negative control group were prepared by combining equal volumes of individual samples. Albumin was depleted from each pooled sample with Pierce™ Albumin Depletion Kit (Thermo Fisher Scientific). Albumin-depleted sample protein concentrations were determined with *DC*™ protein assay (Bio-Rad Laboratories). Each albumin-depleted pooled plasma sample containing 200 μg of total protein was diluted to 300 μl with PBS. Diluted samples were incubated with the resin for 1 hour with gentle agitation. The resin was washed with 400 μl PBS three times. To elute glycoproteins, 75 μl of 0.2 M lactose in PBS was added to the column and incubated with gentle agitation. After 30 minutes, the flow-through was collected. Then 75 μl of 0.2 M lactose in 0.2 M acetic acid was added to the column and incubated with gentle agitation. After 30 minutes, the flow-through was collected and combined with the previous flow-through. The combined eluate was then dialyzed against PBS.

### Mass Spectrometry and Protein Identification

Trypsin digestion was performed on the samples. Samples were reduced (200mm DTT in 50mm bicarbonate) and alkylated (200mM iodoacetamide in 50mm bicarbonate) before trypsin (6 ng/μl, Promega Sequencing grade) was added to a ratio of 1:20. The digestion was done overnight (∼16 hrs) at 37°C and formic acid was then added to adjust the pH to 2-4. The samples were then dried, redissolved in 4% acetonitrile, 0.1% formic acid, and desalted using C18 tips (Thermo Scientific).

Peptides were resolved and ionized by using nano flow HPLC (Easy-nLC 1000, Thermo Scientific) coupled to a Q Exactive Orbitrap mass spectrometer (Thermo Scientific) with an EASY-Spray capillary HPLC column (ES800A, 75um x 15cm, 100Å, 3μm, Thermo Scientific). The mass spectrometer was operated in data-dependent acquisition mode, recording high-accuracy and high-resolution survey orbitrap spectra using external mass calibration, with a resolution of 35,000 and m/z range of 300–1700. The twelve most intense multiply charged ions were sequentially fragmented by using HCD dissociation, and spectra of their fragments were recorded in the orbitrap at a resolution of 17,500; after fragmentation all precursors selected for dissociation were dynamically excluded for 30 s. Data was processed using Proteome Discoverer 1.4 (Thermo Scientific) and the database, Uniprot Human UP000005640, was searched using SEQUEST (Thermo Scientific). Search parameters included a strict false discovery rate (FDR) of 0.01, a relaxed FDR of 0.05, a precursor mass tolerance of 10ppm and a fragment mass tolerance of 0.01Da. Peptides were searched with carbamidomethyl cysteine as a static modification and oxidized methionine and deamidated glutamine and asparagine as dynamic modifications.

Protein quantitation was based on the number of peptide spectral matches (PSM). First, detected proteins (PSM >= 1 in at least one sample) were searched in the online portal of CRAPOme^28^, a database of protein contaminants in proteomic experiments.

CRAPOme outputs a ratio of [Num of Expt. (found/total)] for each query protein. Any protein with a [Num of Expt. (found/total)] > 0.2 is considered a contaminant and removed. To identify non-specifically binding proteins, two tailed student’s t-test was performed between the PSM of the proteins in the triplicates of the pulldown samples and corresponding bead-only controls (PSM_PD_ and PSM_CT_, respectively). Any protein that satisfies 1) average PSM_PD_ <= average PSM_CT_, *or* 2) PSM_PD_ < 2, *or* 3) p-value of the t-test > 0.05, is removed. The remaining proteins are the SNA-enriched proteins and are listed in **Supplementary Table 5** (severe COVID-19) and **Supplementary Table 6** (mild COVID-19).

To identify significantly upregulated proteins in SNA-enriched severe COVID-19 plasma, two tailed student’s t-test was performed between the PSM of the enriched proteins in the triplicates of the severe sample and the mild sample (PSM_severe_ and PSM_mild_, respectively). Any protein that satisfies 1) average PSM_severe_ > average PSM_mild_ *and* 2) p-value of the t-test < 0.05 is considered significantly upregulated in severe COVID-19 plasma and is listed in **Supplementary Table 7**.

### Western Blotting

All steps were conducted at room temperature unless noted otherwise. The column eluate, or the corresponding input (albumin-depleted plasma) containing 20 μg of total protein, was mixed with Laemmli buffer to a final volume of 200 μl. Then 100 μl of each sample was heated at 90°C before resolved by 4-20% SDS-PAGE. Proteins were transferred to a nitrocellulose membrane, which was then stained with Ponceau S. After the total protein stain was erased, the membrane was blocked with a blocking buffer (PBS with 3% (w/v) BSA and 0.05% (v/v) Tween^®^ 20, pH = 7.4) for 1 hour. Then the membrane was incubated with primary antibodies pre-diluted to 1 μg/ml in the blocking buffer for 1 hour. Rabbit anti-human complement C5 antibody (clone# EPR19699-24, Abcam, catalog# ab202039) and mouse anti-human complement C9 antibody (clone# X197, Hycult Biotech, catalog# HM2111) were used for C5 and C9 detection, respectively. The membrane was washed with 0.05% PBST three times for 5 minutes per wash then incubated with secondary antibodies pre-diluted to 0.1 μg/ml in the blocking buffer for 15 minutes. CF™640-conjugated, goat anti-rabbit IgG antibody (Millipore Sigma, catalog# SAB4600399) and IRDye^®^ 800CW-conjugated, goat anti-mouse IgG antibody (LI-COR, catalog# 926-32210) were used for C5 and C9 primary antibody detection, respectively. Finally, the membrane was washed with 0.05% PBST three times for 5 minutes per wash before imaging.

### Immunofluorescence Staining

Formalin-fixed paraffin embedded autopsy tissues was sectioned at 5 μm on Plus Slides (Fisher Scientific, Catalog# 22-042-924) and stored at room temperature prior to use. Sections were probed with the following reagents: Cy5 conjugated *Sambucus nigra* lectin (SNA, Vector Laboratories, catalog# CL-1303); recombinant his-tagged divalent carbohydrate-binding Module 40^29^ (diCBM40, gift from Dr. Anne Imberty, CERMAV-CNRS); recombinant his-tagged Griffithsin^30^ (GRFT, gift from Dr. Weston Struwe, University of Oxford). Lectin fluorescent histochemistry, protein binding fluorescence and chromogenic immunohistochemistry were performed on a Roche Ventana Discovery XT platform using Ventana reagents and detection kits unless otherwise noted. Sections were pre-incubated at 60°C followed by online deparaffinization (Discovery Wash catalog# 950-150). SNA (1.0 mg/ml), diCBM40 (0.8 mg/ml) and GRFT (1.0 mg/ml) were diluted 1:100 in Carbo-Free Blocking Solution (Vector Laboratories catalog# SP-5040, Lot# ZG0630) and incubated for 3 hours at room temperature. diCBM40 and GFRT were detected with Alexa Fluor™ 555 conjugated, mouse anti-6xHis Tag antibody (1.0 mg/ml, Thermo Fisher Scientific, catalog# MA1-21315-A555, Lot# WD326765, RRID: AB_557403) diluted 1:100 in Carbo-Free Blocking Solution and incubated for 60 minutes. Labeled sections were washed in distilled water, counterstained with 100.0 ng/ml DAPI and cover-slipped with Prolong Gold anti-fade media.

### Immunohistochemistry Staining

Single and multiplex-chromogenic immunohistochemistry was performed using unconjugated mouse-anti human Interleukin-6, (clone# OTI3G9, Origene, catalog #TA500067, Lot# VE2990982), unconjugated murine anti-human Vimentin (clone# V9, Ventana Medical Systems, Catalog# 790-2917, Lot# E04396, RRID:AB-2335925) and unconjugated mouse anti-human CD-163 clone MRQ-26 (Ventana Medical Systems catalog# 760-4437, lot# V001041, RRID: AB_2335969), unconjugated murine anti human ST6 beta-galactoside alpha-2,6-sialyltransferase 1, (clone# LN1, Thermo Fisher Scientific, Cat# MA5-11900, Lot# XC3519066A, RRID: AB_10980157^31^), unconjugated murine anti-human Pan-Cytokeratin (PanK, Thermo Fischer Scientific Cat# MA1-82041, Lot# 985542A RRID: AB_2335731), unconjugated mouse anti-human complement C9 antibody, (clone# X197, Hycult Biotech, catalog# HM2111, Lot# 16152M0714-B, RRID: AB_2067596^32^) and unconjugated mouse anti-human terminal complement complex C5b-9, (clone# aEll, Dako Cytomation, catalog# M0777, Lot# 20027911, RRID: AB_2067162^33^). Antibodies were tested and sequence optimized on a composite 30-core tissue microarray containing both normal and tumor tissues. All samples were sectioned at four microns and collected onto Plus microscope slides (Fisher Scientific, catalog# 22-042-924) and stored at room temperature prior to use. Sections for IL6 were pre-incubated at 60°C followed by online deparaffinization (Discovery Wash, catalog# 950-150). Anti-IL6 was diluted 1:50 (20.0 μg/ml) in Ventana Antibody Diluent (catalog# 760-219) and incubated for 5 hours at room temperature. Primary antibody was detected using goat anti-mouse horseradish peroxidase conjugated multimer incubated for 8 minutes. The complex was visualized with 3,3 diaminobenzidine and enhanced with copper sulfate. Slides were washed in distilled water, counterstained with hematoxylin, dehydrated and mounted with permanent media.

Multiplex samples were assayed with a tissue microarray for positive, negative and multiplex crossover control^34^. Multiplex-chromogenic immunohistochemistry was performed on a Ventana Medical Systems Discovery Ultra using Ventana reagents and detection kits unless otherwise specified. In brief, slides for sequential Vimentin-CD163 multiplex were heated at 60°C for 1 hour and deparaffinized on-instrument. Antigen retrieval was performed in CC1 (TRIS-Borate-EDTA, pH = 8.5, Roche, catalog# 950-224) for 32 minutes at 95°C followed by treatment with 3% hydrogen peroxide for 8 min to quench endogenous peroxidase. Anti-Vimentin was applied neat for 20 minutes at 37 °C followed by detection with goat anti-mouse Horseradish Peroxidase conjugated multimer and visualized with purple (TAMRA) chromogen^35^. Slides were denatured in instrument wash buffer (Atlas Antibodies, catalog# 950-330) at 95°C for 32 minutes to strip immunological reagents followed by application of hydrogen peroxide for 8 minutes to quench Horseradish Peroxidase. Anti-CD163 was applied neat and incubated for 60 minutes at 37 °C followed by detection with goat anti-rabbit Horseradish Peroxidase conjugated multimer and visualized with yellow (DABsyl) chromogen^35^. All Slides were washed in distilled water, counterstained with hematoxylin, dehydrated and mounted with permanent media.

Sections for C9 were antigen retrieved using Cell Conditioner 1 (Tris-Borate-EDTA, pH = 8.5, catalog# 950-500) for 20 minutes and C5b9 treated with Protease-3 (catalog# 760-2020) for 12 minutes. Anti-C9 (1.0 μg/ml) and C5b9 (0.045 mg/ml) antibody was diluted 1:100 in TBSA (25mM Tris, 15mM NaCl, 1% BSA, pH = 7.2) and incubated for 3 hours at room temperature. Primary antibody was detected using goat anti-mouse horseradish peroxidase conjugated multimer incubated for 8 minutes. The complex was visualized with 3,3 diaminobenzidine and enhanced with copper sulfate. Slides were washed in distilled water, counterstained with hematoxylin, dehydrated and mounted with permanent media. Negative controls were incubated with diluent only.

### Analysis of Whole Blood RNA-seq Datasets

Publicly available whole blood RNA sequencing data and raw counts analyzed in this work were downloaded from Gene Expression Omnibus (GSE161731) published in a study by McCLain et al.^36^, and analyzed in GraphPad Prism.

### Statistical Analysis

Unpaired, two-tailed Mann-Whitney U-test and Student’s t-test were used for statistical analysis in this study. When statistical analysis is performed, the type of test used and *p*-value annotation are indicated in the corresponding figure captions or in the corresponding subsections of Methods.

## RESULTS

### α2,6-Sialylation is Upregulated in Plasma of Severe COVID-19 Patients

COVID-19 severity is highly variable, ranging from asymptomatic disease to acute respiratory distress syndrome (ARDS) and death^1, 37^. While glycosylation of SARS-CoV-2-specific IgG has been identified as a severity marker^24, 25^, there have been no broader analyses of glycosylation in plasma with regards to COVID-19. Herein, we examined the glycosylation of plasma from a cohort of 71 SARS-CoV-2 positive adults recruited at the University of Alberta Hospital. The majority of samples were collected during the second wave of COVID-19 from October 2020 to January 2021 (**Supplementary Table 1**). Plasma samples were collected at the first hospital visit or at time of a SARS-CoV-2 positive PCR test. We categorized the COVID-19 positive patients into three severity groups: i) patients who were not hospitalized (*mild*, n = 5), ii) patients who were hospitalized but did not need supplemental oxygen (*moderate*, n = 8) and iii) patients who were hospitalized, received supplemental oxygen and/or were in the ICU (*severe*, n = 58). As a negative control, we used pre-pandemic plasma samples from an age- and gender-matched healthy cohort (n = 60) recruited at the University of Georgia (**Fig. 1a**).

**Figure 1.**
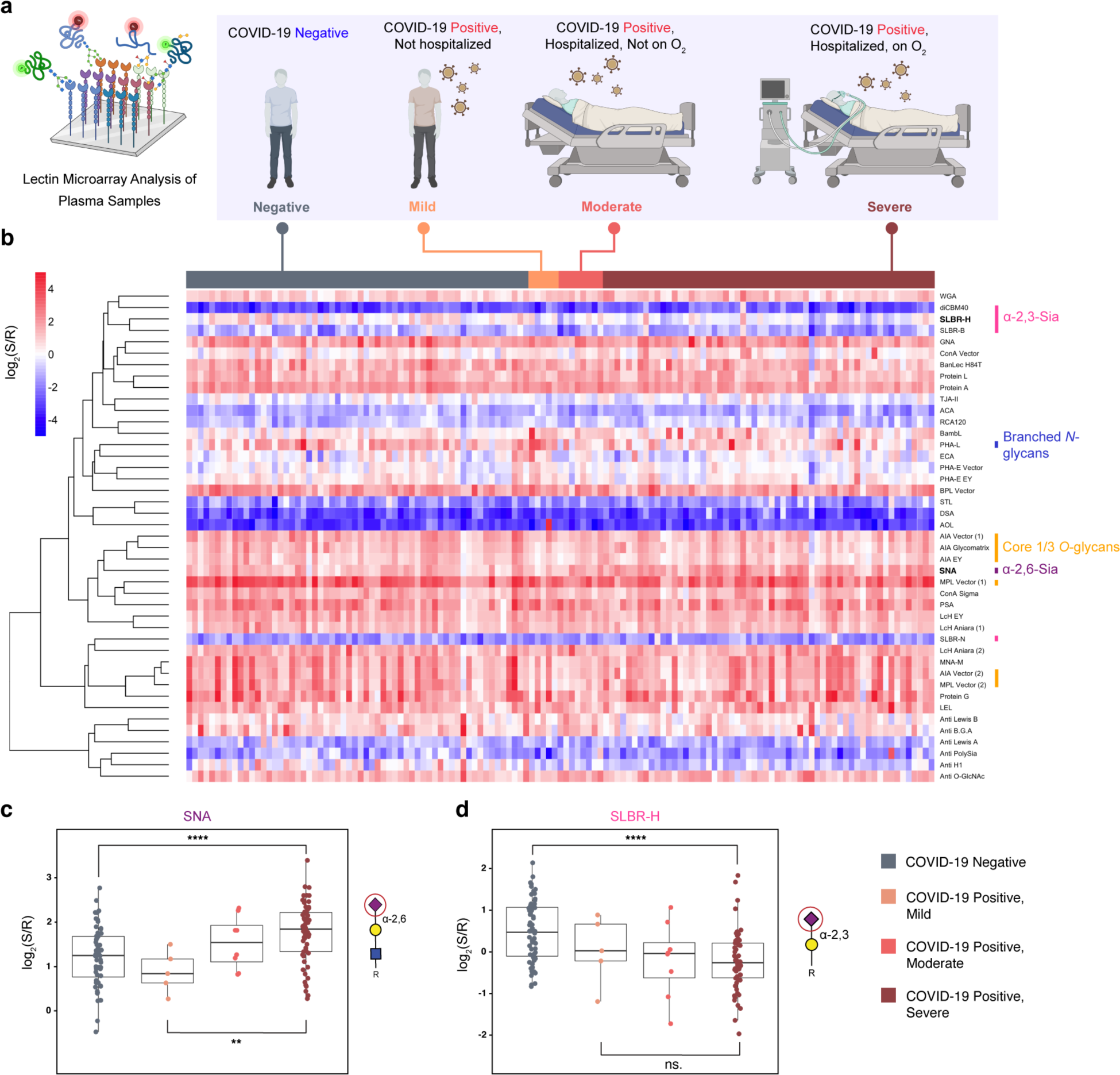
Plasma glycomic profiles of COVID-19 positive and negative cohorts. **(a).** Schematic description of analysis. COVID-19 positive patients were categorized into three groups by disease severity (Mild, Moderate, Severe) and compared to an age and gender-matched control cohort (Negative). Plasma samples were analyzed by lectin microarray. **(b).** Heatmap of lectin microarray data with annotations of rough glycan specificities for select lectins. Columns (patients) are ordered by disease severity as in (a), indicated at the top of the heatmap. **(c).** Boxplot analysis of SNA and SLBR-H binding data by patient group. Glycan ligands for the lectins are shown in the Symbolic Nomenclature for Glycomics (SNFG). Symbols are defined as follows: galactose (yellow •), N-acetylglucosamine (blue), sialic acid (purple ■). Mann-Whitney U test was used to determine *p* values. **: *p* < 0.01; ****: *p* < 0.0001.

To analyze the glycome, we performed our dual-color lectin microarray analysis on the plasma samples (left panel, **Fig. 1a**). Lectin microarray technology utilizes a collection of specific, well-characterized glycan-binding proteins^38^ to evaluate differential glycan expression patterns between sample groups^39^. An advantage of this method is that probes which identify significant changes in the glycome can then be used for further glycoproteomic and histochemical studies. High-throughput glycomic analysis using our established array technology has revealed new roles for glycans in cancer biology, host-pathogen interactions and viral and exosome biogenesis^23, 27, 40–43^. In brief, each sample was fluorescently labelled (S, Alexa Fluor™ 555 labeled) and mixed with an equal amount of an orthogonally labeled reference standard (R, Alexa Fluor™ 647 labeled commercial plasma). Samples were analyzed using in-house fabricated lectin microarrays containing >95 probes (**Supplementary Table 4,Supplementary Table 5**), as previously described^26^. A heatmap of the normalized log_2_ data (S/R) from our COVID-19 positive and control plasma samples is shown in **Fig. 1b**.

Comparison of all COVID-19 positive samples to the controls identified several distinct glycomic changes with infection (**Supplementary Fig. 1**). We observed a significant loss of core 1,3 O-glycans (AIA), α2,3 sialic acid (SLBR-H, SLBR-B) and β1,6 branched N-glycans (PHA-L) and an increase in α2,6 sialic acid (SNA). Closer examination of differences in glycosylation between the severity groups revealed changes in the levels of total α2,6-sialylation of glycoproteins as a function of severity. Patients with severe COVID-19 had significantly higher levels of α2,6-sialylation when compared to either the negative controls or the mild COVID-19 cohort (severe vs. negative: ∼45% increase; severe vs. mild: ∼84% increase, **Fig. 1c**). In contrast, although lower levels of α2,3-sialylation were observed, we did not see severity dependent changes (SLBR-H, SLBR-B, diCBM40, **Fig. 1d**, **Supplementary Fig. 2**). Overall, we observed both infection- and severity-dependent changes in the plasma glycome in COVID-19.

### Post-mortem COVID-19 Patients Exhibited Significant Increase in Sialylation in Select Tissues

COVID-19 can affect multiple organs, causing thromboembolism, kidney injury, damage to the heart, etc^44–47^. To investigate whether changes in plasma are reflective of changes in tissue from COVID-19 patients, we conducted a glycomic analysis of autopsy tissue samples using our lectin microarrays. Tissues were obtained from patients who either succumbed to COVID-19 during the initial phase of the pandemic in New York City (positive, pos) or individuals who died of other causes (negative, neg)^48^. The majority of patients in the cohort had lung pathology. More specific patient characteristics are given in **Supplementary Table 2**. We confirmed the histological integrity and quality of the autopsy tissues (heart, kidney, liver, upper and lower lobe of the lung) by the presence of intact nuclei in hematoxylin and eosin staining (**Fig. 2a**). Heatmap showing significant difference in glycomic assessment of autopsy tissues is shown (**Fig. 2b**). A complete heatmap of lectin microarray data organized by tissue type and COVID status is shown in **Supplementary Fig. 3**.

**Figure 2.**
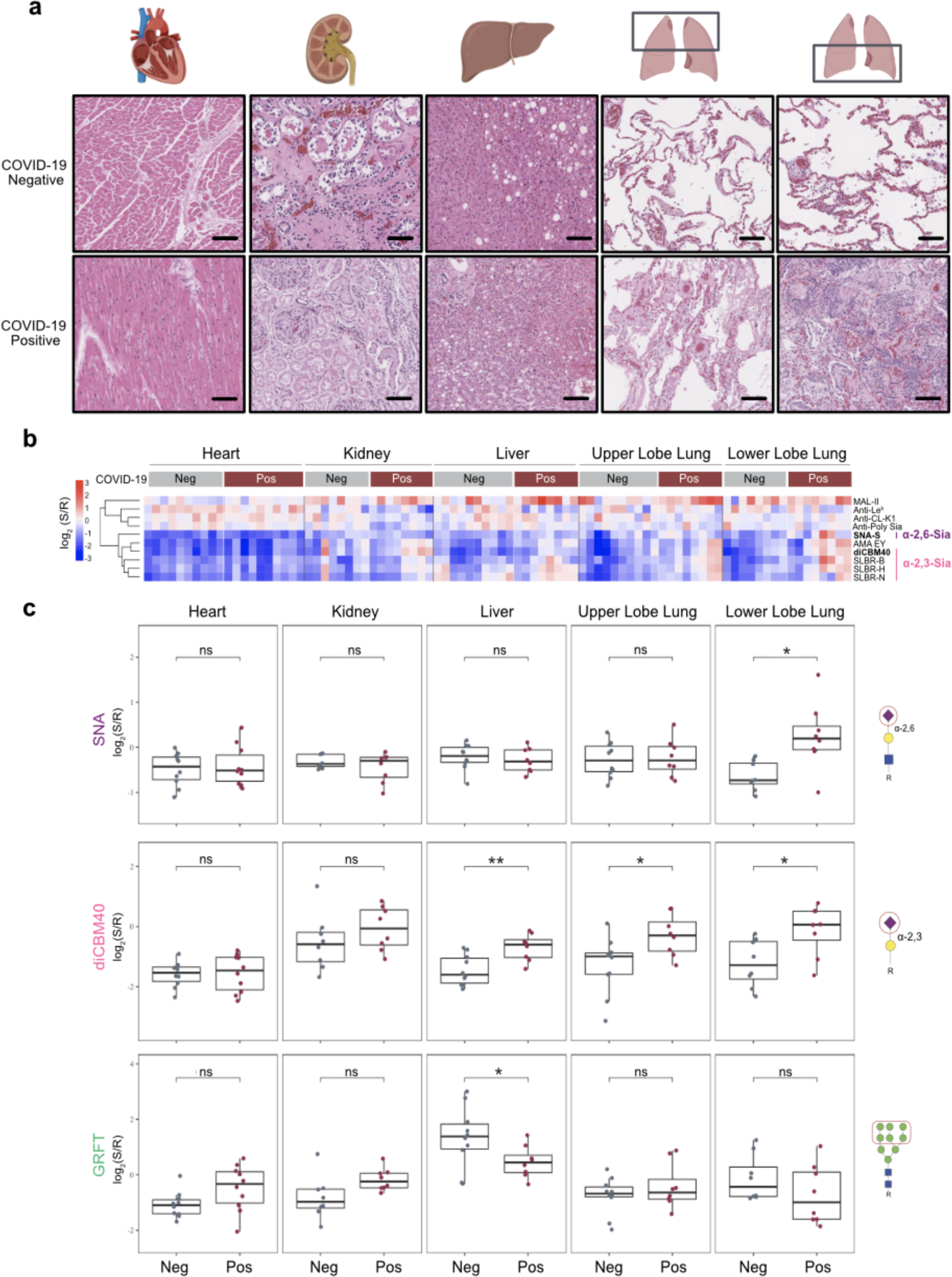
Organ-specific glycan changes are observed in COVID-19 patients. **(a)** Representative images of H&E stains of organs from COVID-19 positive and COVID negative autopsy samples (heart: n=5; kidney: n=4; liver: n=4; upper lobe lung: n=4; lower lobe lung: n=4, 2 samples per patient). Scale bars represent 75 μm. **(b)** Heatmap presenting statistically significant lectins (*p* < 0.05, Student’s t-test) from lectin microarray analysis is shown. Samples from COVID positive (heart: n=5; kidney: n=4; liver: n=4; upper lobe lung: n=4; lower lobe lung: n=4, 2 samples per patient) and COVID negative (heart: n=5; kidney: n=4; liver: n=5; upper lobe lung: n=5; lower lobe lung: n=4, 2 samples per patient) were analyzed. Lectins were hierarchically clustered using Pearson correlation coefficient and average linkage analysis. Median normalized log_2_ ratios (Sample (S) / Reference(R)) were ordered by sample type. Red, log_2_(S) > log_2_(R); blue, log_2_(R) > log_2_(S). **(c)** Boxplot analysis of α-2,6-sialic acids probed by SNA. Boxplot analysis of α-2,3-sialic acids probed by diCBM40. Boxplot analysis of high-mannose probed by GRFT. Data is from analysis shown in (a). COVID-19 positive (maroon), Covid Negative controls (gray). Student’s t-test was used to determine *p*-values. *: *p* < 0.05. Glycans bound by lectins are shown at the right side of boxplots.

Tissue-dependent patterns were observed in our analysis, regardless of mode of death. This follows previous work that showed that the glycome could be used to segregate cell lines by cancer tissue origin and indicates that glycosylation is a marker of tissue type^49^. Within tissues, we observed several COVID-dependent glycan signatures (**Supplementary Fig. 4**). SARS-CoV-2 infections begin in the nasal passages before migrating to the lung, where it causes significant damage that can result in death. It should be noted that the majority of our matched controls exhibited pulmonary pathology upon autopsy (**Supplementary Table 2**). In concordance with our plasma data, an increase in α2,6-sialylation was seen in the lower lobe, but not upper lobe of the lung in COVID-19 patients. In many respiratory illnesses, damage to the lower lobe of the lung is associated with more advanced disease^50^. In contrast to our data from plasma, we also observed a significant increase in α2,3-sialic acid in COVID patients. This increase was seen in both the upper and lower lobes of the lung and in liver tissues (diCBM40, **Fig. 2c**; SLBR-B, SLBR-N and SLBR-H, **Supplementary Fig. 5**). We also see several other changes in glycosylation in the COVID positive cohort that are predominantly in the liver. This group has lower levels of high mannose (GRFT and BanLec H84T, **Fig. 2c**) and higher levels of core 1, 3 O-glycans (MPA, MNA-G, **Supplementary Fig. 6**). Interestingly, higher, albeit non-statistical, levels of high mannose are observed in heart and kidney, both of which have been shown in other systems to undergo MBL2-mediated damage^51, 52^.

We corroborated our lectin microarray findings using lectin fluorescence staining within the same cohort of autopsy specimens. Overall, the staining confirmed lectin microarray findings, including increase in α2,6-sialic acid in the lower lobe of the lungs (SNA, **Fig. 3a**), increase in α2,3-sialic acid in both lung compartments (diCBM40, **Fig. 3b**) and the liver (diCBM40, **Supplementary Fig. 7**) and a decrease in high mannose in the livers of COVID-19 patients (GRFT, **Supplementary Fig. 7**). The upper lobe lung of COVID-19 patients also showed slightly higher α2,6-sialic acid staining (SNA, **Fig. 3a**), although the magnitude of increase was significantly smaller than observed in the lower lobes.

**Figure 3.**
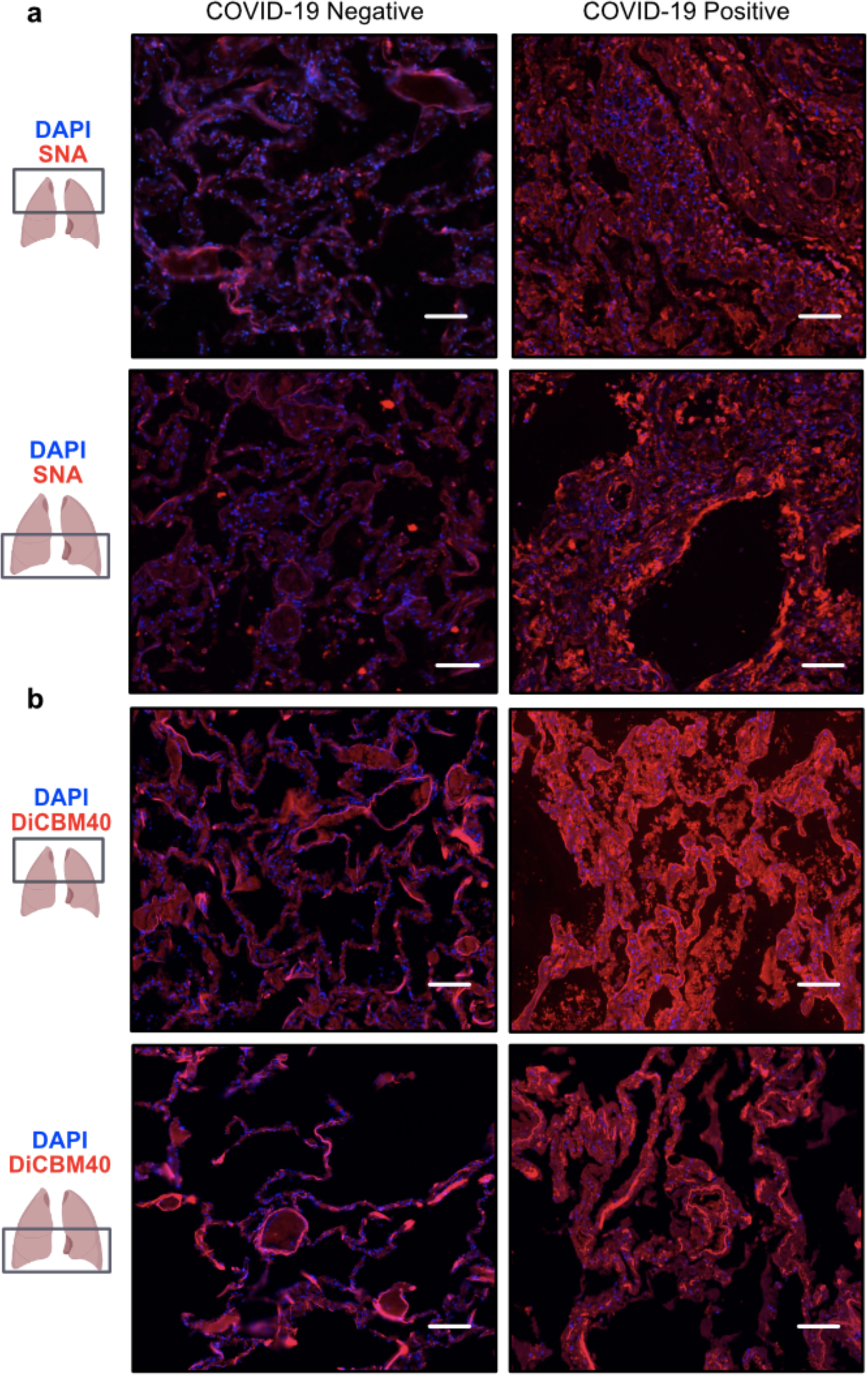
Expression of a2,6 sialic acid (SNA) and a-2,3 sialic acid (diCMB40) in Upper and Lower Lobe of COVID-19 Autopsy Lungs. **(a)** Representative images of IF staining against SNA in the upper lobe (top) or lower lobe (bottom) of the lungs on COVID-19 negative (n = 2) or COVID-19 positive (n = 8) autopsy specimens. Scale bars represent 150 μm. **(b)** Representative images of IF staining against diCBM40 in the upper lobe (top) or lower lobe (bottom) of the lungs on COVID-19 negative (n = 2) or COVID-19 positive (n = 8) autopsy specimens. Scale bars represent 150 μm.

### Complement Proteins Are Differentially Sialylated in Severe COVID-19

Higher levels of α2,6-sialic acid were observed in both our plasma and autopsy cohorts, leading us to wonder whether there was a mechanistic link between these two findings. With this in mind, we conducted α2,6-sialic acid focused glycoproteomic analysis using pooled plasma from the same cohort that we ran glycomic analysis on. In brief, we pooled the plasma of the mild COVID-19 patients and of severe COVID-19 patients and performed SNA pulldown on the two pooled plasma samples. SNA-enriched proteins were then identified using standard mass spectrometry (**Fig. 4a**).

**Figure 4.**
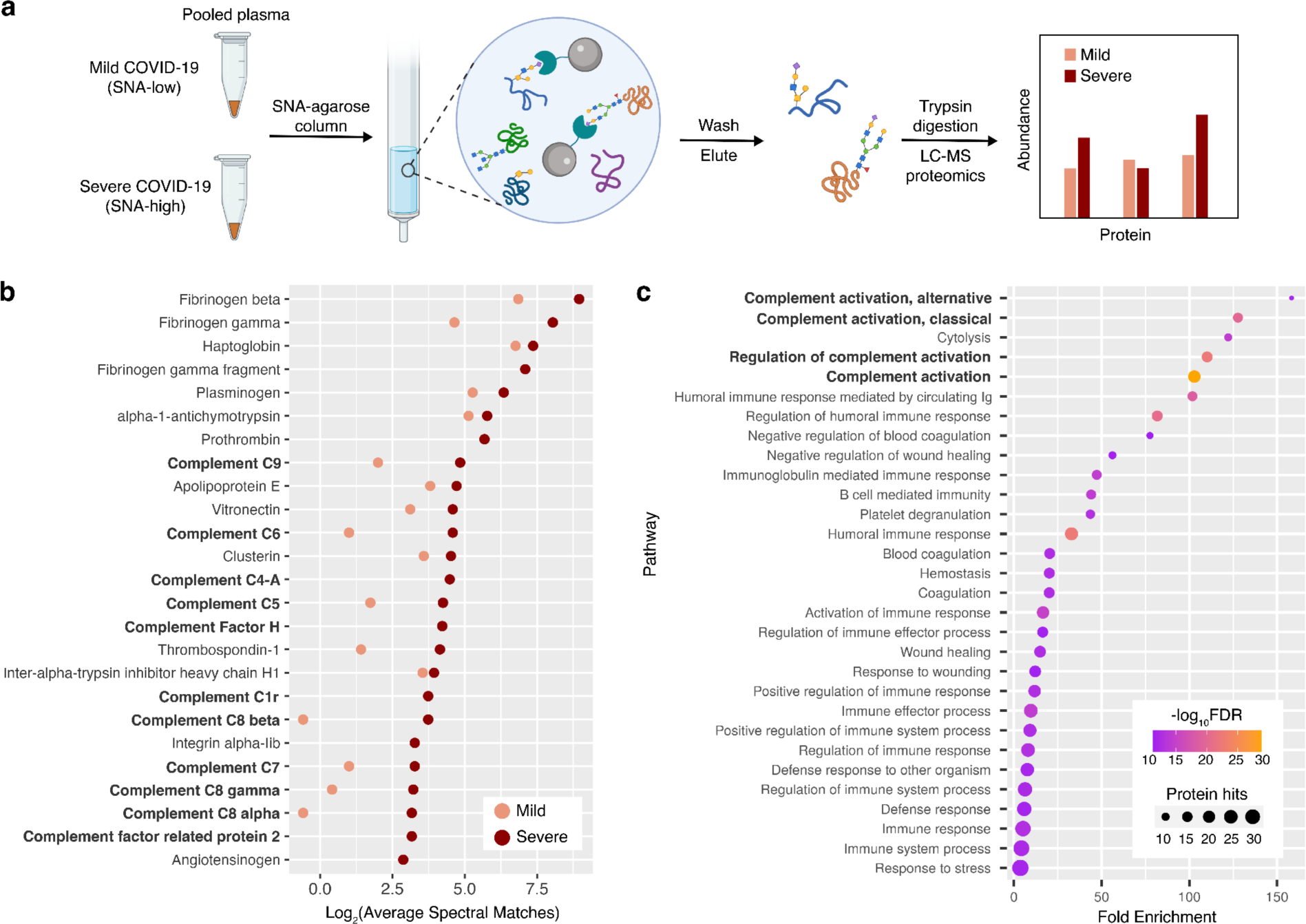
Glycoproteomic analysis of α2,6-sialic acid-containing proteins from mild and severe COVID-19 plasma. **(a)** Scheme of workflow. Mild COVID-19: COVID-19 patients who were not hospitalized. Severe COVID-19: COVID-19 patients who were hospitalized and received supplemental oxygen. **(b)** SNA-reactive glycoproteins significantly enriched the severe COVID-19 plasma compared to mild, and their mass spectrometric abundance profiles (average spectral matches). The top 25 enriched glycoproteins (by abundance) in the severe group, are shown. **(c)** Pathway enrichment analysis of the enriched plasma glycoproteins. Number of protein hits and -log_10_FDR for pathways are indicated.

We identified 77 proteins in the severe COVID-19 group (**Supplementary Table 5)** and 38 in mild (**Supplementary Table 6**), with 29 proteins identified in common between both groups. This was in line with our expectation, as severe COVID-19 patients showed greater plasma α2,6-sialylation. A total of 44 SNA-enriched glycoproteins were significantly higher in abundance in the severe COVID-19 group compared to the mild (**Fig. 4b;Supplementary Table 7**). As expected, we observed many proteins involved in thrombosis and the coagulation cascade, including fibrinogens, plasminogen and prothrombin. This is consistent with findings in autopsy and other severe COVID-19 cohorts, where dysregulated thrombosis is observed^48, 53^. These proteins are often highly abundant in plasma and enrichment could be due to increases in the expression of mediators of coagulation^53–55^. Gene Ontology enrichment analysis using the differentially expressed proteins identified complement system-related pathways as the most significantly enriched (**Fig. 4c**). This implies that α2,6-sialic acid upregulation may be connected to severe COVID-19 via a mechanistic link involving the complement system. We observed many of the downstream complement cascade members in our enriched pool (C5, C6, C7, C8, C9, **Fig. 4b**; **Fig. 5a**).

**Figure 5.**
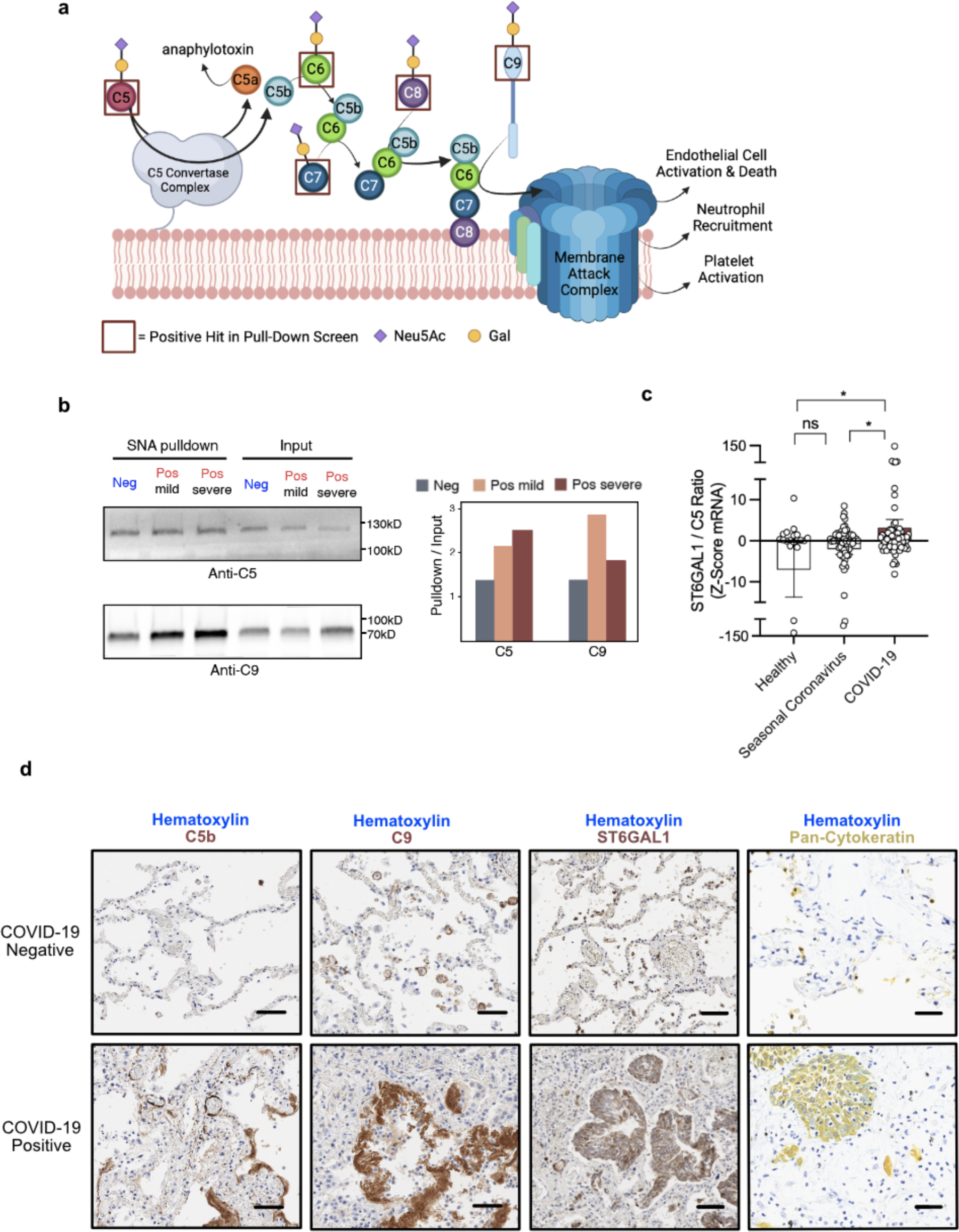
Fraction of α2,6-Sialylated Complement Cascade is enriched in COVID-19 Blood and Lungs. **(a)** Scheme of the later portion of the complement cascade pathway and downstream signaling. Maroon boxes indicate complement proteins that were positive hits in our glycoproteomic analysis in Fig. 4. **(b)** Differential α2,6-sialylation of complement C5 and C9 in three patient groups. Western blot (anti-C5 and anti-C9) of SNA pulldown samples from pooled patient plasma with corresponding input is shown. Intensity ratios of pulldown/input bands are depicted in the bar plot. **(c)** Analysis of individual per patient paired ratio of ST6GAL1 to C5 mRNA expression in whole blood of healthy controls (n = 19) or those with seasonal coronavirus (n = 59), or COVID-19 (n = 46), from publicly available dataset Gene Expression Omnibus: GSE161731. Student’s t-test was used to determine *p*-values. ns: p ≥ 0.05; *: *p* < 0.05. **(d)** Representative images of IHC staining against C5b, C9, ST6GAL1 or Pan-Cytokeratin in the lower lobe of the lungs in COVID-19 negative (n = 2) (top) or COVID-19 positive (n = 8) (bottom) autopsy specimens. Scale bars represent 75 μm.

Activation of the complement system in COVID-19 and its association with severity have been reported by multiple groups^8, 56, 57^. Both C5 and C9 have been associated with severe COVID-19 and contribute to MAC-induced cell death and damage (**Fig. 5a**)^57, 58^. To gain insight into whether we are observing a change in sialylation or complement levels, we performed SNA pulldowns from pooled plasma samples from mild and severe COVID-19 patients and controls and performed Western blot analysis for C5 and C9 (**Fig. 5b**). In the COVID-19 patients, both C5 and C9 have higher levels of α2,6-sialylation, when compared to control, with some evidence of a severity-dependent increase in sialylation for C5. These data suggest an increased fraction of complement is α2,6-sialylated in COVID-19.

Complement proteins can be produced in the liver, immune cells, the endothelia and the epithelia^59, 60^. In COVID-19 patients, we observed aberrant expression of ST6 beta-galactoside alpha-2,6-sialyltransferase 1 (ST6GAL1), the main enzyme responsible for α2,6-sialylation, in the lung epithelia (**Fig. 5d**; **Supplementary Fig. 8**). We also found high levels of expression of ST6GAL1 in the liver of both COVID-19 positive and negative patients, consistent with the known expression patterns for this enzyme (**Supplementary Fig. 8**)^61, 62^. Mining of publicly available RNA-seq data^36^ showed an increased ratio of ST6GAL1 to C5 in the whole blood of patients with COVID-19, when compared to those with seasonal coronavirus infection or to uninfected controls (**Fig. 5c**). Neither ST6GAL1 nor C5 alone showed significantly different levels in COVID-19 when compared to control (**Supplementary Fig. 9**). In aggregate these data suggest that an enhanced fraction of α2,6-sialylated complement proteins, potentially deriving from multiple compartments, is associated with COVID-19 as a disease state.

To examine whether complement activation is observed in our autopsy cohort, we performed immunohistochemical (IHC) staining for C5b and C9. We observed high levels of C5b and C9 deposition in the livers of COVID-19 patients when compared to controls (**Supplementary Fig. 8**). We also observed extensive deposition of C5b and C9 in the lungs of COVID-19 patients (**Fig. 5d**). In the lower lobe of the lung, staining for both complement proteins and α2,6-sialic acid is concentrated at the edges of the airway barrier, in line with our observation that complement proteins themselves are sialylated. The complement cascade can have many downstream effects^8, 9^. IL-6, which is strongly associated with COVID-19 mortality, is both a promoter of and enhanced by complement activation^63, 64^. In line with this, staining of our autopsy cohort showed increased IL-6 levels in the lungs in COVID-19 patients compared to controls (**Supplementary Fig. 10**). In multiple pathogenic diseases, the complement cascade is profibrotic, acting directly on myeloid cells (e.g., macrophages) and fibroblasts alike^65–70^. Complement is also known to recruit myeloid cells to sites of tissue injury. Recent literature suggests that in COVID-19, there is enhanced pulmonary fibrosis mediated by CD163^+^ macrophages that exhibit fibroblast-like features^71^. In our cohort, we observed a drastic enrichment of this cell type in the lower lobe of COVID-19 positive lungs (**Supplementary Fig. 10**). Our cohort showed pathological outcomes characteristic of the complement hyperactivation, suggesting a possible link between sialylation and complement mediated tissue damage in COVID-19.

## DISCUSSION

Glycosylation has multifaceted roles in immunity and host-response to pathogens^14, 17, 72^. Recognition of glycans help determine self vs. non-self and can trigger immune activation via both the innate and adaptive immune system. In influenza, severity of disease was found to be associated with levels of high mannose and the innate immune lectin MBL2^23, 42, 73^. In SARS-CoV-2 infection, antibody glycosylation has been studied as a marker of severity. Antibodies to the spike protein were altered in severe patients, with lower fucosylation and sialylation observed^24, 25^. This has potential consequences for effector function^15, 22^. However, such studies have focused on a single protein type (IgG). To date there has been no work on the systemic glycomic response to SARS-CoV-2 infection in plasma and no analysis of infected tissues.

Herein, we performed high-throughput analysis of plasma and autopsy sample glycosylation from COVID-19 patients using our lectin microarray technology. Our analysis revealed plasma α2,6-sialic acid as a marker of severity. This modification is known to increase the half-life of select proteins, including IgG^74, 75^. We also observed higher levels of α2,6-sialic acid in the lower lobe of the lungs in patients who have died from COVID-19. In previous studies, CT scans showed that lower lobe involvement and consolidation is common in COVID-19 patients^76, 77^. In the lower lobe, staining for α2,6-sialic acid appeared strongest at the barrier of blood-gas exchange.

Glycoproteomic analysis of α2,6-sialylated proteins from plasma showed enrichment in members of the complement cascade. The complement cascade is a proteolytic cascade culminating in the formation of the membrane attack complex (MAC, **Fig. 5a**). Activated through both innate and adaptive immune mechanisms, it stimulates multiple immune responses including myeloid cell mobilization, cytokine release, cell damage, platelet activation and the coagulation cascade^3–6^. Hyperactivation of the complement cascade is recognized as an emerging therapeutic target for COVID-19^8, 12, 78^. In line with this, we observed high levels of staining for complement proteins C5 and C9 in COVID-19 autopsy samples. Of note, in the lower lobe lung the staining for complement also localized to the barrier of blood-gas exchange.

In plasma, we found that the fraction of α2,6-sialylated C5 and C9 in severe COVID-19 patients is significantly upregulated (**Fig. 5b**). The increased pool of sialylated complement most likely derived from augmented expression of ST6GAL1, the main enzyme responsible for α2,6-sialylation. We observed upregulation of this enzyme in the lung (**Fig. 5d**; **Supplementary Figure 8**). In addition, analysis of previous work found higher relative levels of this enzyme in the blood in COVID-19 patients (**Fig. 5c**). In concordance with our findings, several studies have also shown upregulation of ST6GAL1 in lung epithelium, liver and immune cells in COVID-19^79–81^. Collectively, the data suggests α2,6-sialylation plays a role in the immune response to COVID-19.

Previous studies have shown that almost all complement proteins can bear sialylated glycans^82–84^. Factor H, which inhibits the cascade, binds α2,3 sialylation on host-cells, a critical aspect of self-recognition^85^. However, the functional significance of sialylation on complement proteins remains poorly understood. Several works point to a role for glycosylation on complement proteins in controlling immune function. Gerard et al. showed the de-N-glycosylated form of C5a desArg, a de-arginated proinflammatory anaphylatoxin derived from C5a, was 10- to 12-fold more potent^86^. In contrast, Konterman and Rauterberg showed de-N-glycosylation of C9 dampened the cell lysis activity of C9^87^. Glycosylation can also play a role in controlling both serum half-life and resistance to proteolytic cleavage, which is of particular importance to this cascade^88, 89^. The α2,6-sialylation may be increasing half-life, prolonging the cell-mediated damage from the cascade. There may also be other effects of α2,6-sialylation on complement biology that have yet to be discovered. In general, glycosylation as an aspect of complement has long been ignored. As we seek to develop therapeutic approaches to reverse the detrimental responses from the complement cascade observed in COVID-19, we will need to understand the functional impact of α2,6 sialylation and other glycans on complement.

## Data Availability

Lectin microarray data for plasma samples and autopsy samples are available at Synapse.org (doi: 10.7303/syn27791795).
All other data produced in the present work are contained in the manuscript

https://doi.org/10.7303/syn27791795

## ACKNOWLEDGEMENTS

The authors thank the Alberta Proteomics and Mass Spectrometry Facility for providing mass spectrometry services. The NYULH Center for Biospecimen Research and Development, Histology and Immunohistochemistry Laboratory (RRID:SCR_018304), is supported in part by the Laura and Isaac Perlmutter Cancer Center Support Grant (NIH/NCI P30CA016087). This project has been funded by the National Institute of Allergy and Infectious Diseases, a component of the NIH, Department of Health and Human Services, under contract 75N93019C00052 (T.M.R and L.K.M.) and the Canada Excellence Research Chair Program (L.K.M.). Some graphical contents were created with biorender.com.

## AUTHOR CONTRIBUTIONS

L.K.M. designed the study; D.J., A.H., T.A., B.R., M.A.C., E.K. and A.R. collected the samples; R.Q., E.K., S.C., L.C. and B.Z. performed the experiments; R.Q., E.K., S.C. and L.K.M. analysed the data; R.Q., E.K., S.C. and L.K.M. wrote the paper. D.B.S., T.M.R. and L.K.M. supervised the study.

## DATA AVAILABILITY

Lectin microarray data for plasma samples and autopsy samples are available at Synapse.org (doi: 10.7303/syn27791795).

