## Supplemental Information for "α2,6-Sialylation is Upregulated in Severe COVID-19 Implicating the Complement Cascade"

---

‡ Co-first authorship.

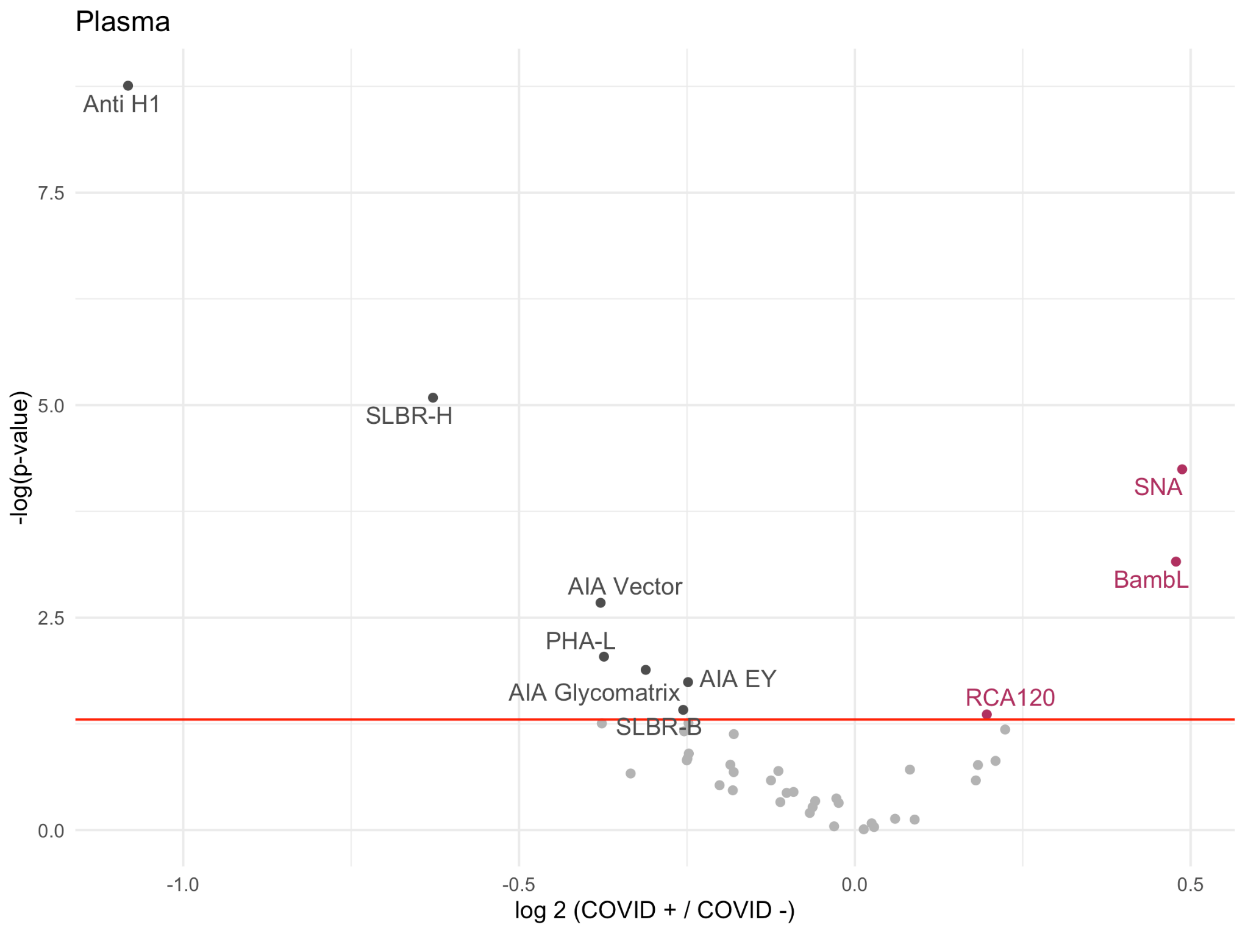


**Supplementary Figure 1. Volcano plot comparing lectin microarray data for COVID-19 positive patients (n = 71) and negative controls (n = 60).** Probes whose binding is statistically significantly decreased in the COVID-19 cohort when compared to controls are on the left of the volcano plot (dark grey). Probes whose binding is increased are on the right (maroon). The red line indicates $p < 0.05$ by the Mann–Whitney U test.

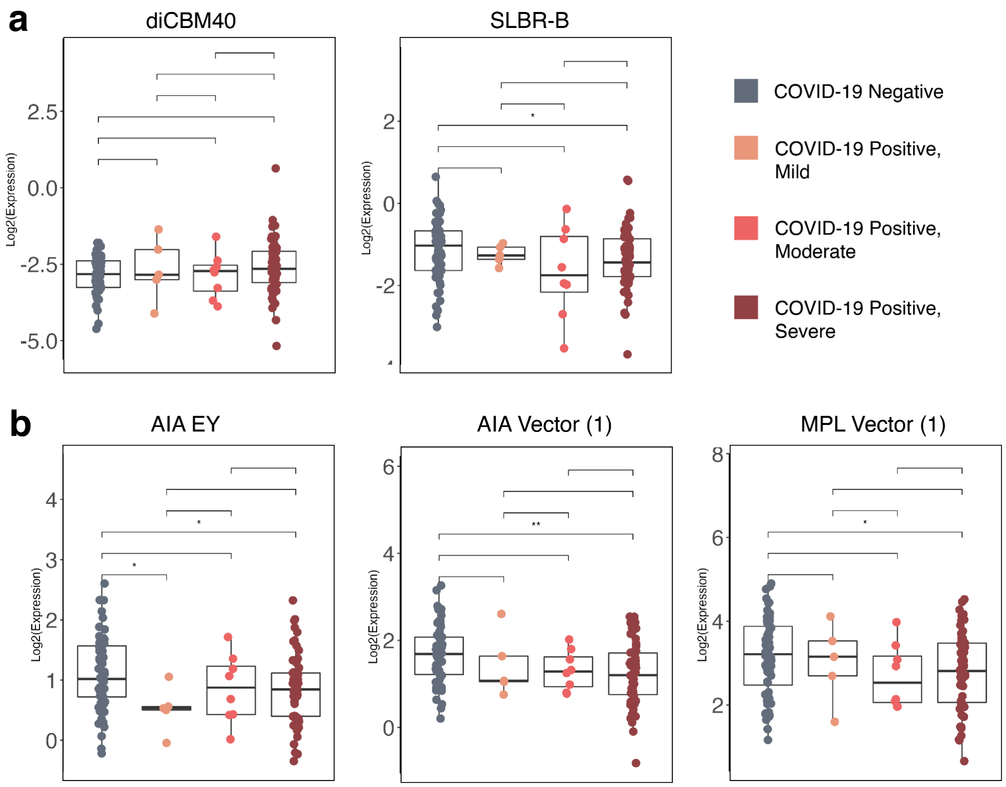


**Supplementary Figure 2**: **Boxplot analysis of (a) α-2,3-sialic acid binding lectins and (b) core 1/3 *O*-glycan binding lectins between patient groups.** Mann-Whitney U test was used to determine *p* values. *: $p < 0.05$; **: $p < 0.01$.

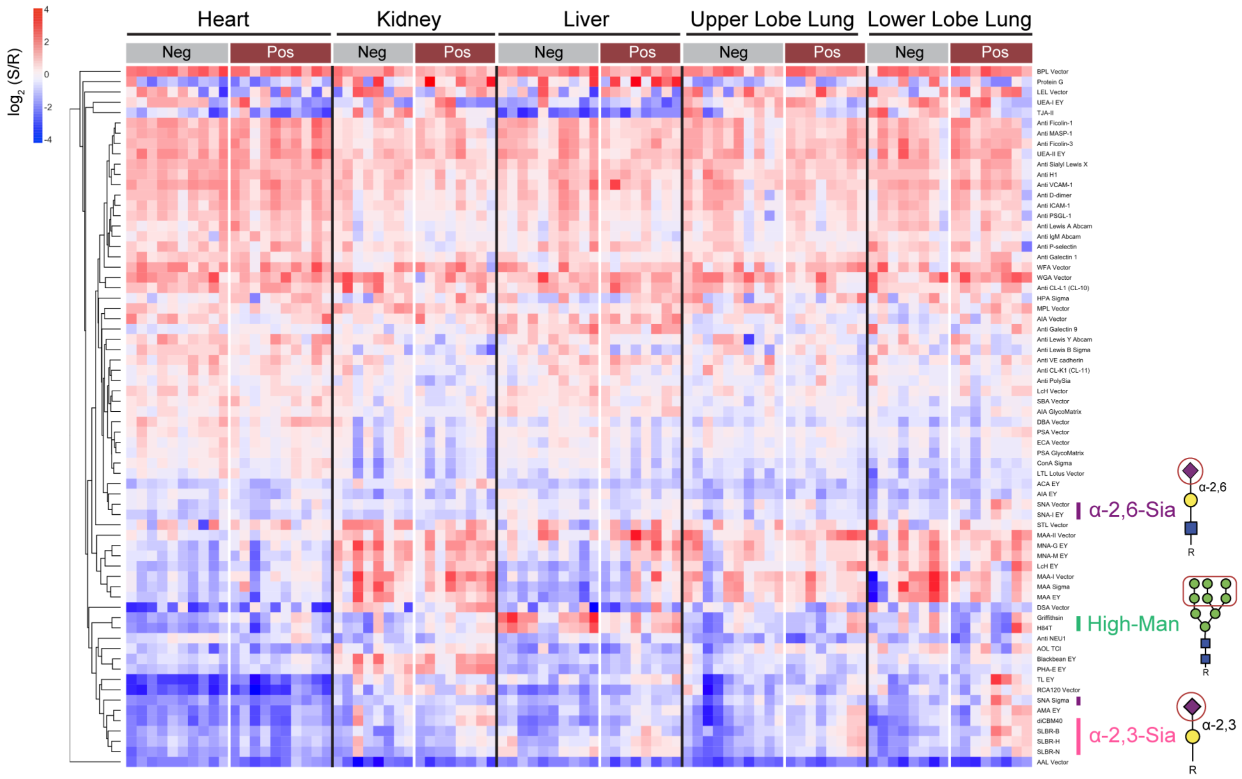


**Supplementary Figure 3. Multi-organ glycomic analysis of COVID-19 deceased patients.** Heatmap of lectin microarray data with the complete list of lectins. Samples from organs of patients who died of COVID (heart: n=5; kidney: n=4; liver: n=4; upper lobe lung: n=4; lower lobe lung: n=4, 2 samples per patient) or other causes (heart: n=5; kidney: n=4; liver: n=5; upper lobe lung: n=5; lower lobe lung: n=4, 2 samples per patient) were analyzed. Median normalized log2 ratios (Sample (S)/Reference(R)) were ordered by organ and COVID status. Red, $\log_2(S) > \log_2(R)$; blue, $\log_2(R) > \log_2(S)$. Lectins binding α2,3-sialosides (pink), α2,6-sialosides (purple) and high-mannose (bright green) are highlighted to the right of the heatmap.

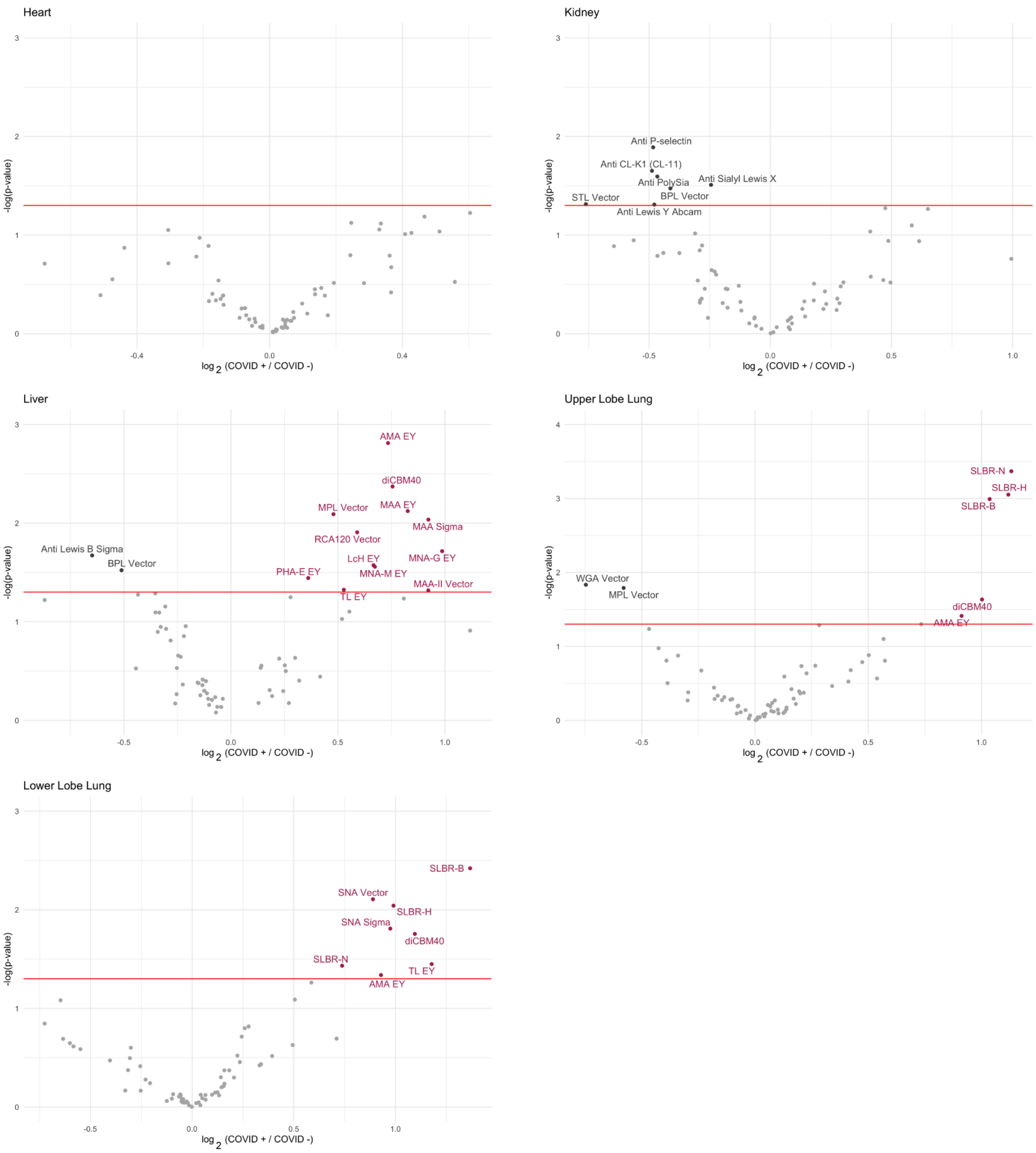


**Supplementary Figure 4. Volcano plot depicting glycan changes of individual organ of COVID-19 deceased patients.** Probes whose binding is statistically significantly decreased in the COVID-19 cohort when compared to controls are on the left of the volcano plot (dark grey). Probes whose binding is increased are on the right (maroon). The red line indicates $p < 0.05$ by Student's t-test.

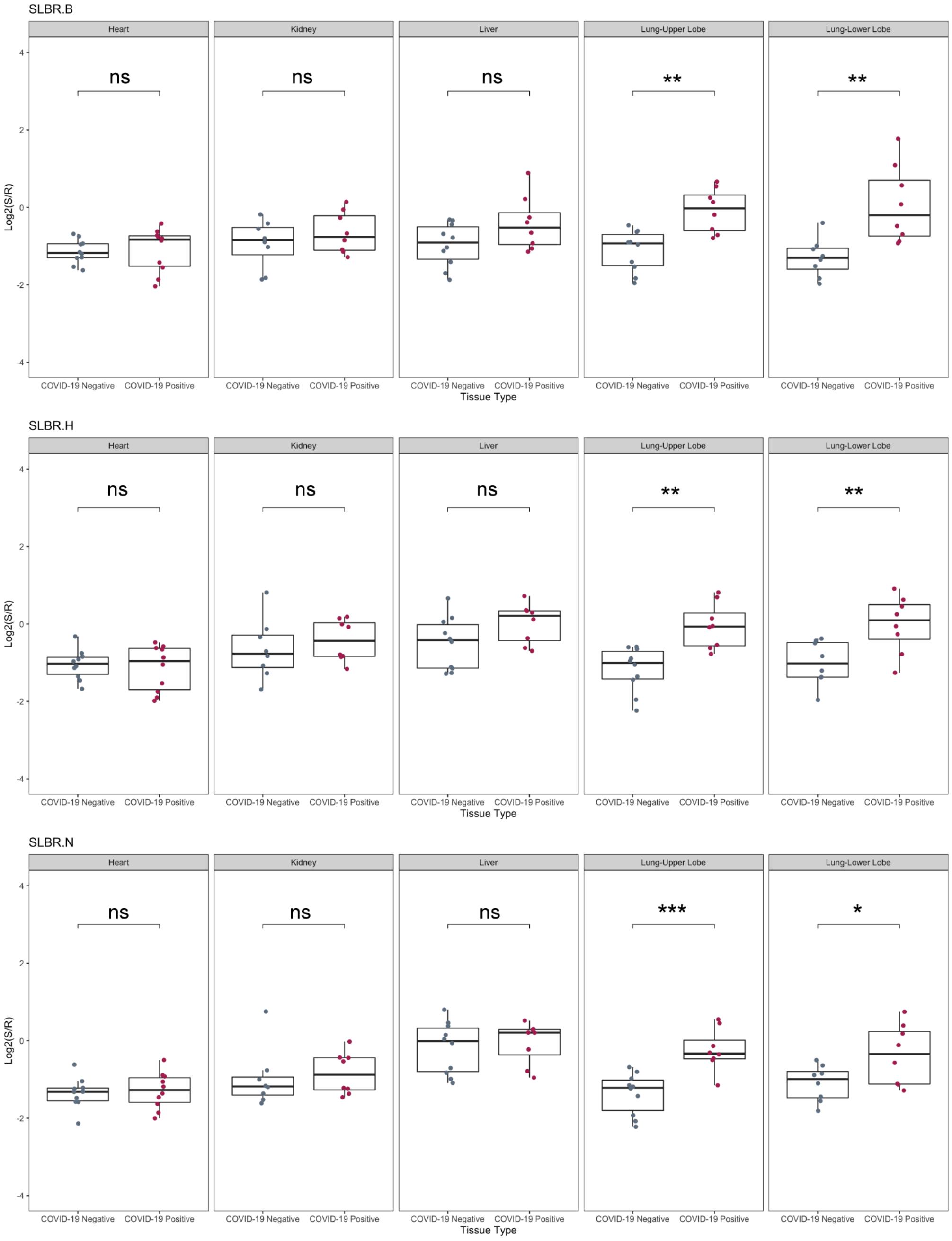


**Supplementary Figure 5. Boxplot analysis of α2,3-sialic acids by SLBR-B, SLBR-H, SLBR-N.** Organs (from left to right): heart, kidney, liver, upper lobe of lung and lower lobe of lung from patients deceased from COVID-19 (maroon), and from other incidents as negative controls (gray). P-value is derived from Student's t-test and red * indicates $p < 0.05$.

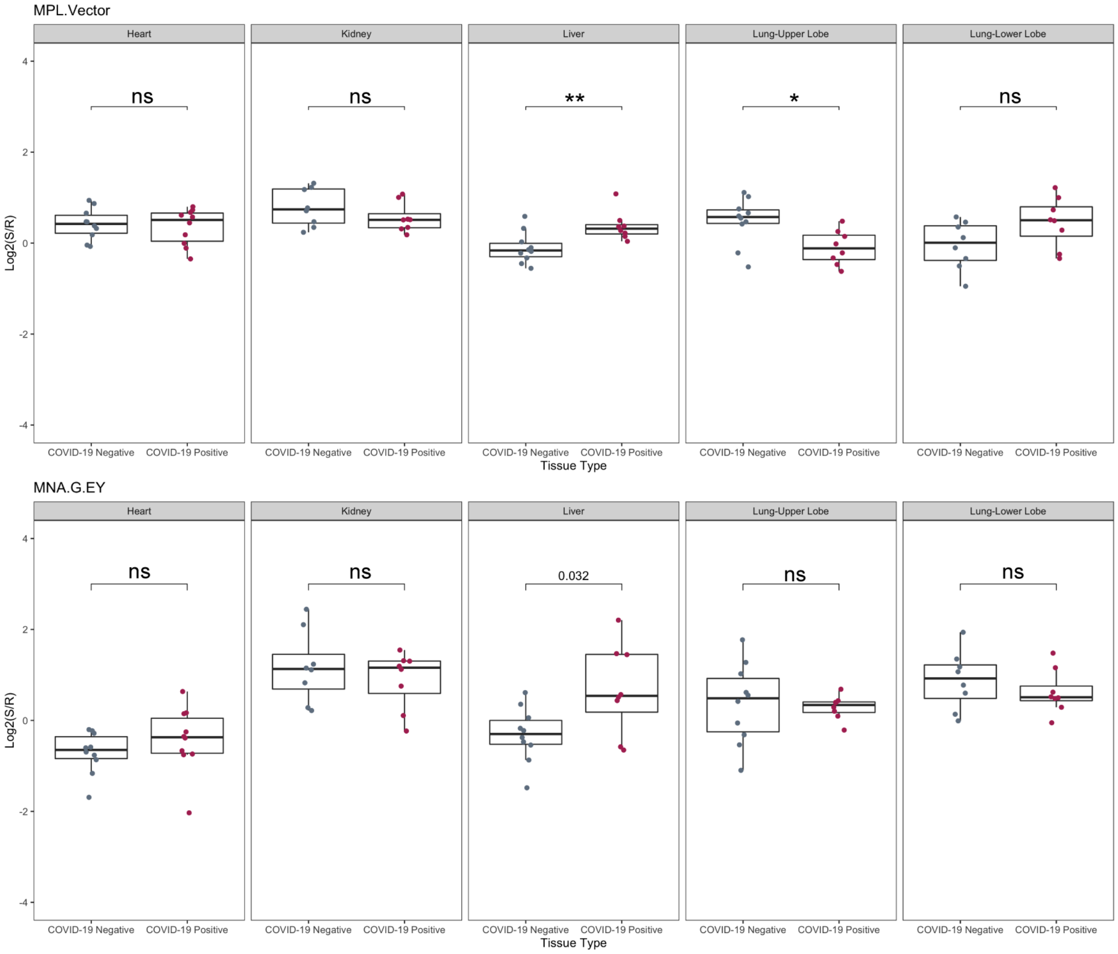


**Supplementary Figure 6. Boxplot analysis of core 1/3 *O*-linked glycans by MPL and MNA-G.** Organs (from left to right): heart, kidney, liver, upper lobe of lung and lower lobe of lung from patients deceased from COVID-19 (maroon), and from other incidents as negative controls (gray). P-value is derived from Student's t-test and red * indicates $p < 0.05$.

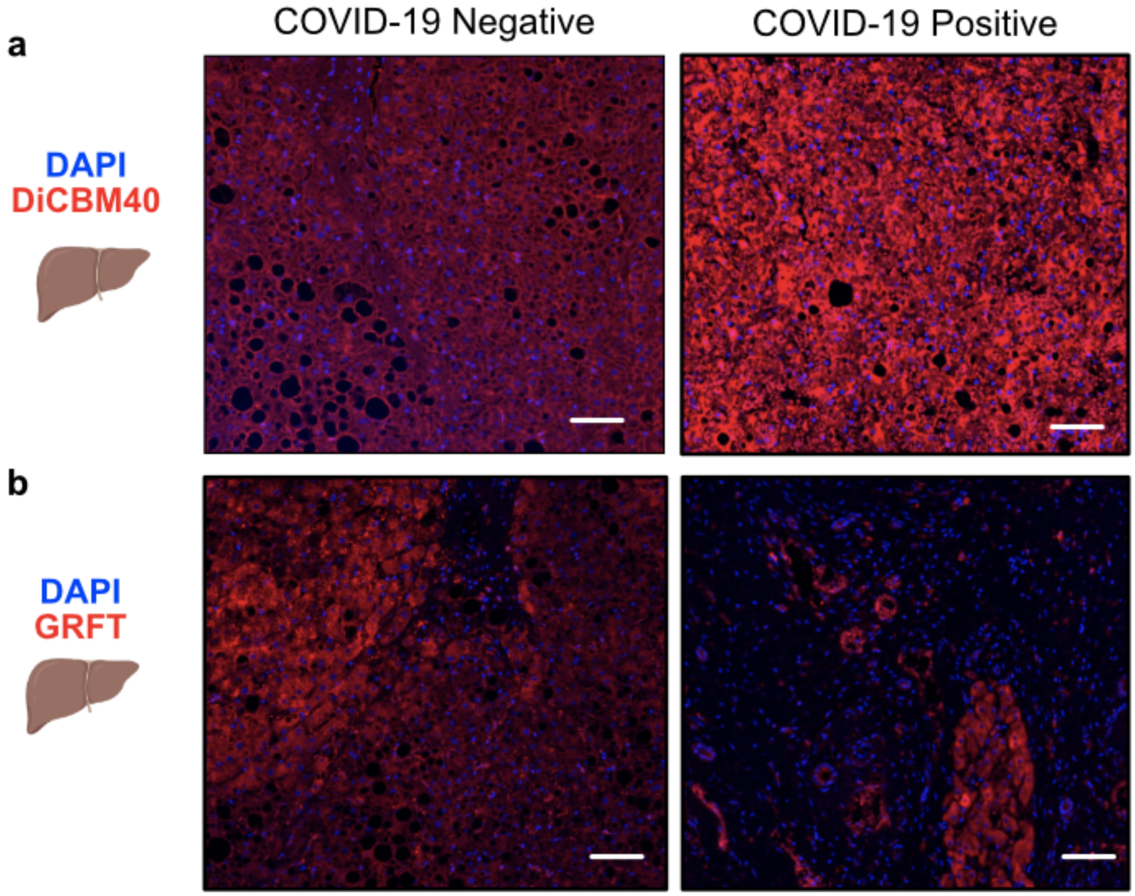


**Supplementary Figure 7. Expression of Select Lectins in COVID-19 Autopsy Livers. (a)** Representative images of IF staining against diCBM40 in the liver in COVID-19 negative (n = 2) or COVID-19 positive (n = 8) autopsy specimens. Scale bars represent 200 µm. **(b)** Representative images of IF staining against GRFT in the liver in COVID-19 negative (n = 2) or COVID-19 positive (n = 8) autopsy specimens. Scale bars represent 200 µm.

.

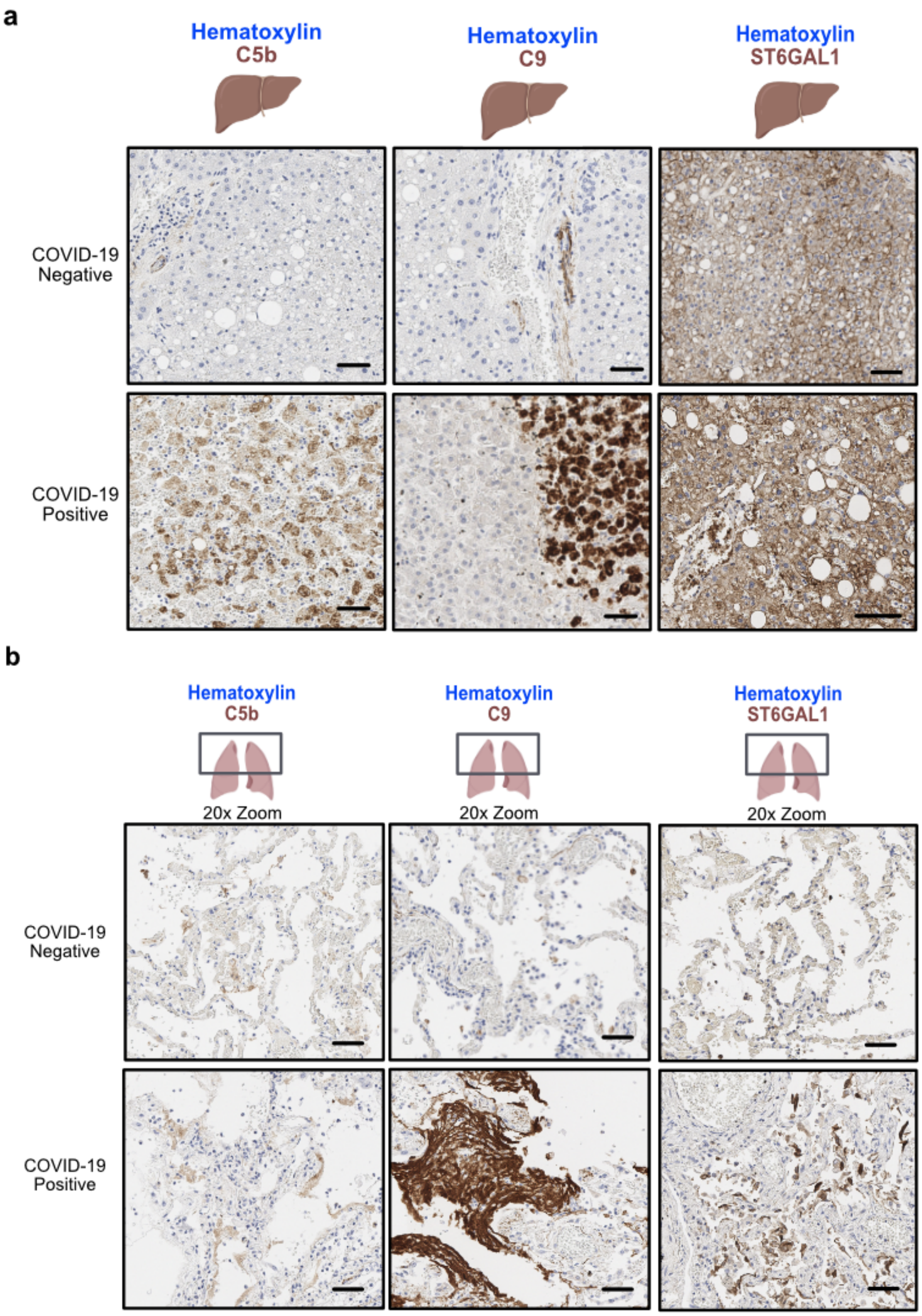


**Supplemental Figure 8. Complement and ST6GAL1 Expression in COVID-19 Autopsy Lungs and Livers. (a)** Representative images of IHC staining against complement C5b (left), C9 (middle) and ST6GAL1 (right) in the liver of COVID-19 negative (n = 2) or COVID-19 positive (n = 8) autopsy specimens. Scale bars represent 100 µm. **(b)** Representative images of IHC staining against complement C5b (left), C9 (middle) and ST6GAL1 (right) in the upper lobe of COVID-19 negative (n = 2) or COVID-19 positive (n = 8) autopsy specimens.

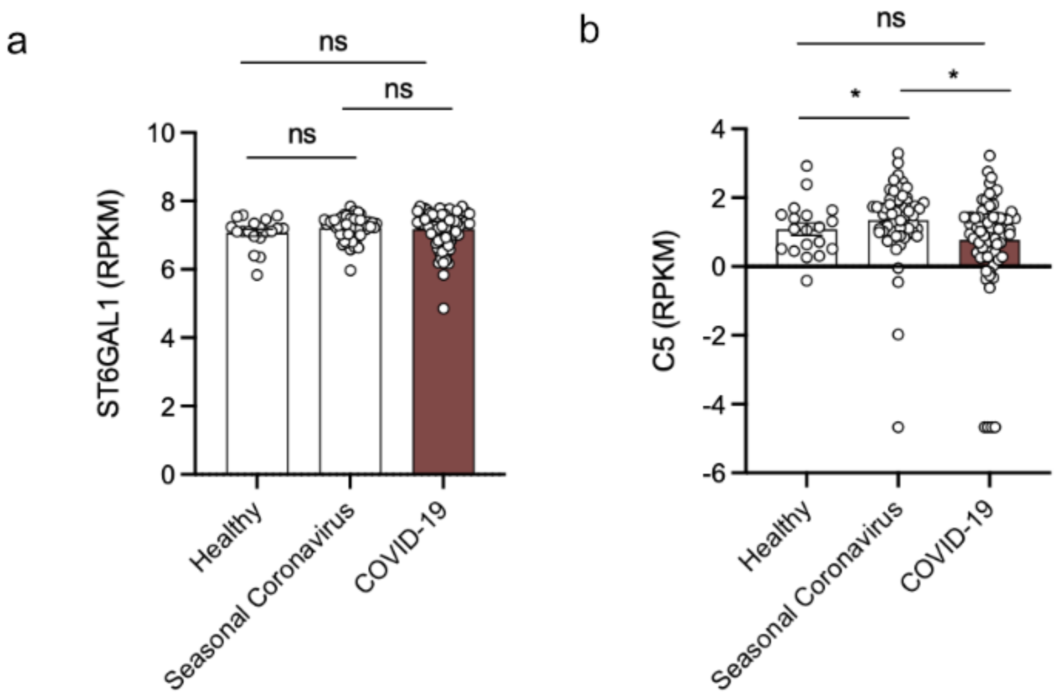


**Supplemental Figure 9. ST6GAL1 and C5 Expression on Whole Blood RNA Sequencing in COVID-19 and Seasonal Coronavirus.** ST6GAL1 **(a)** and C5 **(b)** mRNA expression in whole blood of healthy controls (n = 19) or those with seasonal coronavirus (n = 59), or COVID-19 (n = 46), from Gene Expression Omnibus: GSE161731.

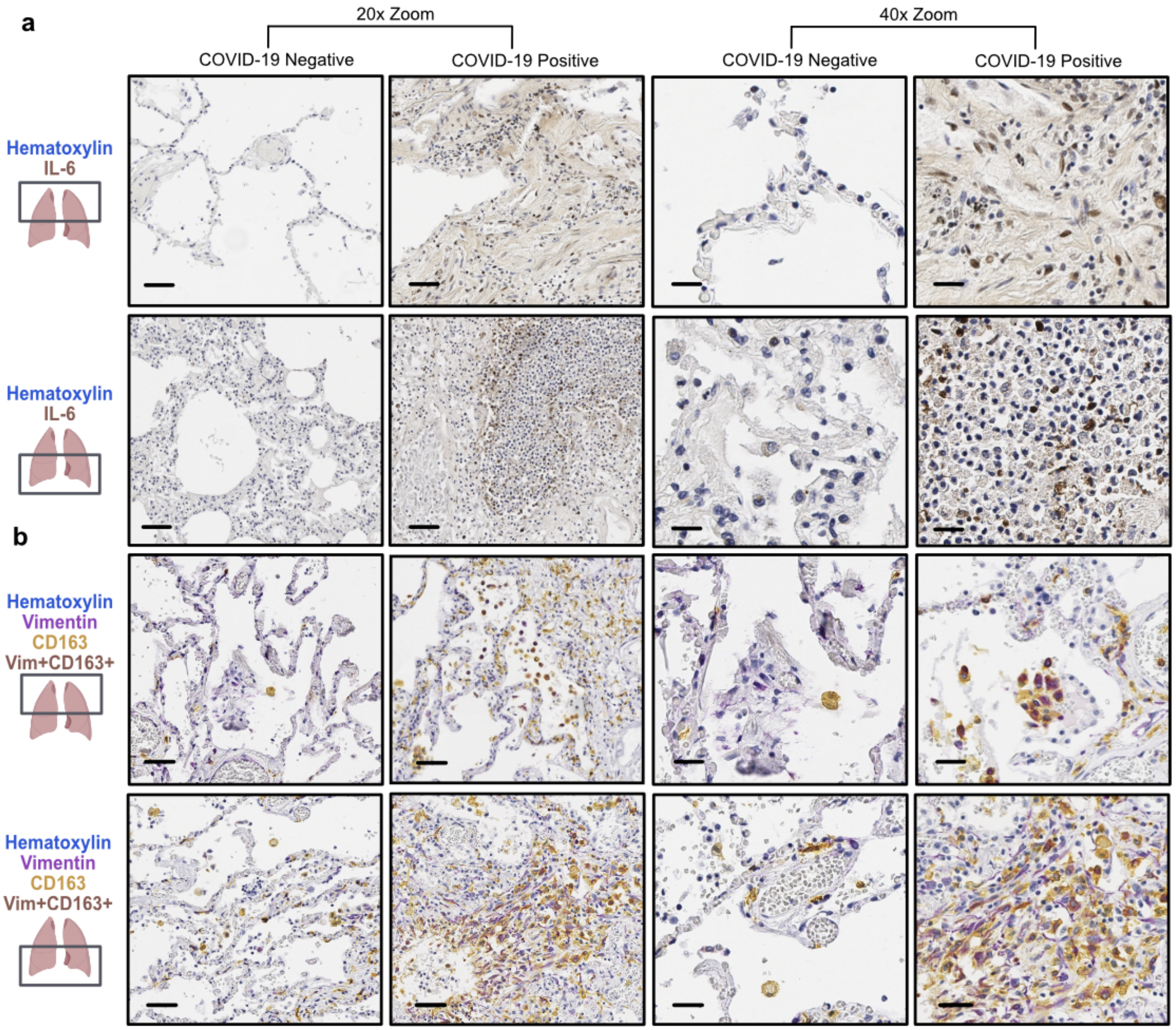


**Supplemental Figure 10. IL-6+ and CD163+Vimentin+ Cells Enriched in Lower Lobe of Lungs of COVID-19 Positive Autopsy Samples. (a)** Representative images of IHC staining against IL-6 in the lower lobe of the lung of COVID-19 negative (n = 2) or COVID-19 positive (n = 8) autopsy specimens. Scale bars represent 100 m on left, and 50 µm on right, throughout. **(b)** Representative images of multi-chromogenic IHC staining against vimentin (purple), CD163 (yellow), and hematoxylin (blue) showing double positive cells in brown in the lower lobe of the lung in COVID-19 negative (n = 2) or COVID-19 positive (n = 8) autopsy specimens (left). Scale bars represent 100 µm on left, and 50 µm on right, throughout.

**Supplementary Table 1. Descriptive Characteristics of Cohorts for COVID-19 Negative and COVID-19 Positive Plasma Samples**

| | **COVID-19 Positive (n=71)** | **COVID-19 Negative (n=60)** |
|---|---|---|
| **Male/Female (ratio)** | 39/30 (1.30) | 35/25 (1.40) |
| **Median Age in Years (IQR)** | 61 (50-73) | 61 (50-71) |
| **Collection Date Ranges in YYYY/MM-YYYY/MM (n)** | 2020/07-2020/09 (2)<br>2020/10-2021/01 (64)<br>2021/02-2021/04 (5) | 2019/09-2019/10 (60) |

Note:
(1) The gender information of 2 participants of the COVID-19 positive cohort is not available.
(2) The age information of 2 participants of the COVID-19 positive cohort is not available.

**Supplementary Table 2. Patient Characteristics for COVID-19 Negative and COVID-19 Positive Autopsy Samples**

| Patient | Age: | Gender: | COVID -19 Status: | Diagnoses: | Pulmonary Pathology (Y/N) |
|---|---|---|---|---|---|
| 1 | 50-60 | Male | Negative | -DAD; pleural effusion<br>-MI; hypertrophy<br>-Cirrhosis, Hep C+ | Y |
| 2 | 60-70 | Male | Negative | -Subarachnoid Hemorrhage, internal carotid artery dissection | N |
| 3 | 0-10 | Female | Negative | -GBM<br>-Aspiration pneumonia (Staph Aureus)<br>-Cardiomegaly | Y |
| 4 | 70-80 | Female | Negative | -DAD<br>-MI; coronary artery disease | Y |
| 5 | 70-80 | Female | Negative | -CHF<br>-intra-abdominal hemorrhage<br>-CKD; pneumonia | Y |
| 6 | 70-80 | Female | Negative | -DAD; pulmonary infarcts, lung congestion** | Y |
| 7 | 40-50 | Male | Negative | -Cor pumonale due to pericarditis and hemorrhage;<br>-pulmonary emboli | Y |
| 8 | 60-70 | Male | Negative | -pleural fluid<br>-Coronary artery disease<br>-hepatomegaly | N |
| 9 | 70-80 | Male | Negative | -Disseminated intravascular coagulation to sepsis<br>-DAD; cardiomegaly | Y |
| 9 | 70-80 | Male | Negative | -Hypovolemic shock<br>-CHF<br>-pulmonary edema with pleural effusions | Y |
| 10 | 60-70 | Female | Positive | -COPD, lung cancer | Y |
| 11 | 50-60 | Female | Positive | N/A | Y |
| 12 | 70-80 | Female | Positive | -Coronary artery disease, adrenal adenoma<br>-COPD | Y |
| 13 | 60-70 | Female | Positive | -Renal cell carcinoma, partial nephrectomy<br>-DAD | Y |
| 14 | 60-70 | Male | Positive | -CAD<br>-acute MI | Y |
| 15 | 50-60 | Female | Positive | -Hyperthyroidism<br>-DAD | Y |
| 16 | 40-50 | Male | Positive | -DAD<br>-Acute Kidney Injury | Y |
| 17 | 60-70 | Male | Positive | -Cirrhosis, CKD<br>-PVD | Y |

Key: DAD = Diffuse Alveolar Damage; MI = myocardial ischemia; CHF = congestive heart failure; CKD = chronic kidney disease; CAD = coronary artery disease
**Autopsy Limited to Lungs Only.

**Supplementary Table 3. Microarray Lectin List for Plasma Analysis**

| Lectin/Antibody | Supplier | Species of Origin | Printing concentration. (mg/ml) | Inhibiting Sugar* |
|---|---|---|---|---|
| ACA | EY | *Amaranthus caudatus* | 2.0 | GlcNAc |
| AIA | EY | *Artocarpus integrifolia* | 2.0 | Gal |
| AIA | Vector (lot#1) | *Artocarpus integrifolia* | 2.0 | Gal |
| AIA v2 | Vector (lot#2) | *Artocarpus integrifolia* | 2.0 | Gal |
| AIA | GlycoMatrix | *Artocarpus integrifolia* | 1.5 | Fuc |
| Anti Lewis A | Abcam | / | as received | / |
| Anti Lewis B | Sigma | / | as received | / |
| Anti O-GlcNAc | Thermo Fisher | / | as received | / |
| Anti PolySia | Absolute Antibody | / | as received | / |
| Anti PSGL-1 | Sigma | / | as received | / |
| Anti Sialyl Lewis X | GeneTex | / | as received | / |
| Anti VCAM-1 | Sigma | / | as received | / |
| Anti VE cadherin | Thermo Fisher | / | as received | / |
| AOL | TCI America | *Aspergillus oryzae* | 2.0 | Fuc |
| BamBL | *Burkholderia cepacia* | expressed in-house | 1.5 | Fuc |
| BanLec H84T | UAlberta | *Musa paradisiaca* | 1.0 | Man |
| BPL | Vector | *Bauhinia purpurea* | 2.0 | Gal |
| ConA | Sigma | *Canavalia ensiformis* | 2.0 | Man |
| ConA | Vector | *Canavalia ensiformis* | 2.0 | Man |
| diCBM40 | UAlberta | *Clostridium perfringens* | 1.5 | Lac |
| DSA | Vector | *Datura stramonium* | 2.0 | Lac |
| ECA | Vector | *Erythrina cristagalli* | 2.0 | GlcNAc |
| LcH | Aniara (lot #1) | *Lens culinaris* | 1.5 | Man |
| LcH | Aniara (lot #2) | *Lens culinaris* | 1.5 | Man |
| LcH | EY | *Lens culinaris* | 2.0 | Man |
| LEL | Vector | *Lycopersicon esculentum* | 2.0 | GlcNAc |
| MNA-M | EY | *Morniga M* | 2.0 | Man |
| MPL | Vector | *Machura pomifera* | 2.0 | Gal |
| PHA-E | EY | *Phaseolus vulgaris* | 2.0 | GlcNAc |
| PHA-E | Vector | *Phaseolus vulgaris* | 2.0 | GlcNAc |
| PHA-L | EY | *Phaseolus vulgaris* | 2.0 | GlcNAc |
| Protein A | Thermo Fisher | / | 0.5 | / |
| Protein G | Thermo Fisher | / | 1.0 | / |
| PSA | Vector | *Pisum sativum* | 2.0 | Man |
| RCA120 | Vector | *Ricinus communis* | 2.0 | Gal |
| SLBR-B | UAlberta | *Streptococcus gordonii* | 2.3 | Lac |
| SLBR-H | UAlberta | *Streptococcus gordonii* | 2.0 | Lac |
| SLBR-N | UAlberta | *Streptococcus gordonii* | 2.0 | Lac |
| SNA | Vector | *Sambucus nigra* | 2.0 | Lac |
| STL | Vector | *Solanum tuberosum* | 2.0 | GlcNAc |
| TJA-II | Aniara | *Trichosanthes jopanica* | 2.0 | Lac |
| WGA | Vector | *Triticum vulgare* | 2.0 | GlcNAc |

*Inhibiting sugars: Man, mannose; Gal, galactose; Fuc, fucose; GlcNAc, N-acetylglucosamine; Lac, lactose.

### Supplementary Table 4. Microarray Lectin List for Autopsy Sample Analysis

| Lectin/Antibody | Supplier | Species of Origin | Printing concentration. (mg/ml) | Inhibiting Sugar* |
|---|---|---|---|---|
| AAL | Vector | *Aleuria aurantia* | 1.5 | Fuc |
| ACA | EY | *Amaranthus caudatus* | 2.0 | GlcNAc |
| AIA | EY | *Artocarpus integrifolia* | 2.0 | Gal |
| AIA | Vector | *Artocarpus integrifolia* | 2.0 | Gal |
| AIA | GlycoMatrix | *Artocarpus integrifolia* | 1.5 | Fuc |
| AMA | EY | *Arum maculatum* | 2.0 | Man |
| Anti CL-K1 (CL-11) | LSBio | / | as received | / |
| Anti CL-L1 (CL-10) | LSBio | / | as received | / |
| Anti D-dimer | Thermo Fisher | / | as received | / |
| Anti Ficolin-1 | LSBio | / | as received | / |
| Anti Ficolin-3 | LSBio | / | as received | / |
| Anti Galectin 1 | Abcam | / | as received | / |
| Anti Galectin 9 | R&D Systems | / | as received | / |
| Anti H1 | Thermo Fisher | / | as received | / |
| Anti ICAM-1 | Thermo Fisher | / | as received | / |
| Anti IgM | Abcam | / | as received | / |
| Anti Lewis A | Abcam | / | as received | / |
| Anti Lewis B | Sigma | / | as received | / |
| Anti Lewis Y | Abcam | / | as received | / |
| Anti MASP-1 | Abcam | / | as received | / |
| Anti NEU1 | Thermo Fisher | / | as received | / |
| Anti P-selectin | Thermo Fisher | / | as received | / |
| Anti PolySia | Absolute Antibody | / | as received | / |
| Anti PSGL-1 | Sigma | / | as received | / |
| Anti Sialyl Lewis X | GeneTex | / | as received | / |
| Anti VCAM-1 | Sigma | / | as received | / |
| Anti VE cadherin | Thermo Fisher | / | as received | / |
| AOL | TCI America | *Aspergillus oryzae* | 2.0 | Fuc |
| BanLec H84T | UAlberta | *Musa paradisiaca* | 1.0 | Man |
| Blackbean | EY | *Phaseolus vulgaris* | 2.0 | Lac |
| BPL | Vector | *Bauhinia purpurea* | 2.0 | Gal |
| ConA | Sigma | *Canavalia ensiformis* | 2.0 | Man |
| DBA | Vector | *Dolichos Biflorus* | 2.0 | Gal |
| diCBM40 | UAlberta | *Clostridium perfringens* | 1.5 | Lac |
| DSA | Vector | *Datura stramonium* | 2.0 | Lac |
| ECA | Vector | *Erythrina cristagalli* | 2.0 | GlcNAc |
| Griffithsin | UAlberta | *Griffithsia* | 1.7 | Man |
| LcH | Vector | *Lens culinaris* | 1.5 | Man |
| LcH | EY | *Lens culinaris* | 2.0 | Man |
| LEL | Vector | *Lycopersicon esculentum* | 2.0 | GlcNAc |
| LTL Lotus | Vector | *Lotus tetragonolobus* | 2.0 | Fuc |
| MAA | Sigma | *Maachia amurensis* | 2.0 | Lac |
| MAA-I | Vector | *Maachia amurensis* | 2.0 | Lac |
| MAA-II | Vector | *Maachia amurensis* | 1.0 | Lac |
| MNA-G | EY | *Morniga G* | 2.0 | Gal |
| MNA-M | EY | *Morniga M* | 2.0 | Man |
| MPL | Vector | *Machura pomifera* | 2.0 | Gal |
| PHA-E | EY | *Phaseolus vulgaris* | 2.0 | GlcNAc |
| Protein G | Thermo Fisher | / | 1.0 | / |
| PSA | Vector | *Pisum sativum* | 2.0 | Man |
| PSA | GlycoMatrix | *Pisum sativum* | 2.0 | Man |
| PTA Gal | EY | *Psophocarpus tetragonolobus* | 2.0 | Gal |
| PTA GalNAc | EY | *Psophocarpus tetragonolobus* | 2.0 | Gal |
| RCA120 | Vector | *Ricinus communis* | 2.0 | Gal |
| SBA | Vector | *Glycine max* | 2.0 | Gal |

| | | | | |
|---|---|---|---|---|
| SLBR-B | UAlberta | *Streptococcus gordonii* | 2.3 | Lac |
| SLBR-H | UAlberta | *Streptococcus gordonii* | 2.0 | Lac |
| SLBR-N | UAlberta | *Streptococcus gordonii* | 2.0 | Lac |
| SNA | Vector | *Sambucus nigra* | 2.0 | Lac |
| SNA | Sigma | *Sambucus nigra* | 2.0 | Lac |
| SNA-I | EY | *Sambucus nigra* | 2.0 | Lac |
| STL | Vector | *Solanum tuberosum* | 2.0 | GlcNAc |
| TJA-II | Aniara | *Trichosanthes jopanica* | 2.0 | Lac |
| TL | EY | *Tulipa sp.* | 2.0 | GlcNAc |
| UEA-I | EY | *Ulex europaeus* | 2.0 | Fuc |
| UEA-II | EY | *Ulex europaeus* | 2.0 | GlcNAc |
| WFA | Vector | *Wisteria floribunda* | 2.0 | Gal |
| WGA | Vector | *Triticum vulgare* | 2.0 | GlcNAc |

*****Inhibiting sugars: Man, mannose; Gal, galactose; Fuc, fucose; GlcNAc, N-acetylglucosamine; Lac, lactose.

**Supplementary Table 5**. Proteins in SNA-enriched Severe COVID-19 Plasma

| Protein Name | UniProt Entry Name | Accession | Average Number of Spectral Matches |
|---|---|---|---|
| Fibrinogen beta chain | FIBB_HUMAN | P02675 | 496 |
| Fibrinogen alpha chain | FIBA_HUMAN | P02671 | 292 |
| Fibrinogen gamma chain | FIBG_HUMAN | P02679 | 264 |
| Complement C3 | CO3_HUMAN | P01024 | 236 |
| Apolipoprotein B-100 | APOB_HUMAN | P04114 | 193 |
| Alpha-1-antitrypsin | A1AT_HUMAN | P01009 | 157 |
| Alpha-2-macroglobulin | A2MG_HUMAN | P01023 | 141 |
| Fibrinogen gamma chain (Fragment) | C9JPQ9_HUMAN | C9JPQ9 | 136 |
| Complement factor H | CFAH_HUMAN | P08603 | 125 |
| Plasminogen | PLMN_HUMAN | P00747 | 81 |
| Hemopexin | HEMO_HUMAN | P02790 | 73 |
| Immunoglobulin kappa constant | IGKC_HUMAN | P01834 | 67 |
| Alpha-1-antichymotrypsin | AACT_HUMAN | P01011 | 55 |
| Prothrombin | THRB_HUMAN | P00734 | 51 |
| Immunoglobulin heavy constant mu | IGHM_HUMAN | P01871 | 45 |
| Inter-alpha-trypsin inhibitor heavy chain H4 | ITIH4_HUMAN | Q14624 | 45 |
| Apolipoprotein A-IV | APOA4_HUMAN | P06727 | 39 |
| Immunoglobulin heavy constant alpha 2 (Fragment) | A0A0G2JMB2_HUMAN | A0A0G2JMB2 | 36 |
| C3/C5 convertase | B4E1Z4_HUMAN | B4E1Z4 | 36 |
| Fibronectin | FINC_HUMAN | P02751 | 36 |
| Complement component C9 | CO9_HUMAN | P02748 | 29 |
| Apolipoprotein E | APOE_HUMAN | P02649 | 26 |
| Vitronectin | VTNC_HUMAN | P04004 | 24 |
| Complement component C6 | CO6_HUMAN | P13671 | 24 |
| Clusterin | CLUS_HUMAN | P10909 | 23 |
| Alpha-1B-glycoprotein | A1BG_HUMAN | P04217 | 23 |
| Complement C4-A | CO4A_HUMAN | P0C0L4 | 22 |
| Leucine-rich alpha-2-glycoprotein | A2GL_HUMAN | P02750 | 20 |
| Complement C5 | CO5_HUMAN | P01031 | 19 |
| Complement factor H | A0A0D9SG88_HUMAN | A0A0D9SG88 | 19 |
| Gelsolin | GELS_HUMAN | P06396 | 18 |
| Thrombospondin-1 | TSP1_HUMAN | P07996 | 18 |
| Lipopolysaccharide-binding protein | LBP_HUMAN | P18428 | 17 |
| Hemoglobin subunit alpha | HBA_HUMAN | P69905 | 17 |
| Alpha-2-HS-glycoprotein | FETUA_HUMAN | P02765 | 16 |
| Centrosomal protein of 290 kDa (Fragment) | A0A5K1VW81_HUMAN | A0A5K1VW81 | 16 |
| Inter-alpha-trypsin inhibitor heavy chain H1 | ITIH1_HUMAN | P19827 | 15 |
| Complement subcomponent C1r | A0A3B3ISR2_HUMAN | A0A3B3ISR2 | 13 |
| Complement component 8 subunit beta | F5GY80_HUMAN | F5GY80 | 13 |
| Alpha-2-antiplasmin | A2AP_HUMAN | P08697 | 13 |
| Immunoglobulin lambda constant 2 | IGLC2_HUMAN | P0DOY2 | 13 |
| Complement factor I | G3XAM2_HUMAN | G3XAM2 | 12 |
| Pregnancy zone protein | PZP_HUMAN | P20742 | 12 |
| Serum amyloid P-component | SAMP_HUMAN | P02743 | 11 |
| Integrin alpha-IIb | ITA2B_HUMAN | P08514 | 10 |
| Complement component C7 | CO7_HUMAN | P10643 | 10 |
| Complement component C8 gamma chain | CO8G_HUMAN | P07360 | 9 |
| Uncharacterized protein C9orf43 | CI043_HUMAN | Q8TAL5 | 9 |
| Platelet factor 4 | PLF4_HUMAN | P02776 | 9 |
| Complement component C8 alpha chain | CO8A_HUMAN | P07357 | 9 |
| Complement factor H-related protein 2 | V9GYE7_HUMAN | V9GYE7 | 9 |
| Fermitin family homolog 3 | URP2_HUMAN | Q86UX7 | 9 |
| Afamin | AFAM_HUMAN | P43652 | 8 |
| Angiotensinogen | A0A7P0T9S6_HUMAN | A0A7P0T9S6 | 7 |

| | | | |
|---|---|---|---|
| C-reactive protein | CRP_HUMAN | P02741 | 6 |
| Carboxypeptidase N catalytic chain | CBPN_HUMAN | P15169 | 6 |
| Ficolin-3 | FCN3_HUMAN | O75636 | 5 |
| Pigment epithelium-derived factor | PEDF_HUMAN | P36955 | 5 |
| Plasma kallikrein | KLKB1_HUMAN | P03952 | 5 |
| Pleckstrin | PLEK_HUMAN | P08567 | 5 |
| Apolipoprotein L1 | APOL1_HUMAN | O14791 | 4 |
| Plasma serine protease inhibitor | IPSP_HUMAN | P05154 | 4 |
| Thyroxine-binding globulin | THBG_HUMAN | P05543 | 4 |
| Coagulation factor V | FA5_HUMAN | P12259 | 3 |
| 2-phospho-D-glycerate hydro-lyase | A0A2R8Y6G6_HUMAN | A0A2R8Y6G6 | 3 |
| Potassium voltage-gated channel subfamily H member 1 | A0A0S1TJ81_HUMAN | A0A0S1TJ81 | 3 |
| N-acetylmuramoyl-L-alanine amidase | PGRP2_HUMAN | Q96PD5 | 3 |
| Immunoglobulin lambda-like polypeptide 5 | IGLL5_HUMAN | B9A064 | 3 |
| Integrin beta (Fragment) | H3BM21_HUMAN | H3BM21 | 3 |
| Apolipoprotein C-II | APOC2_HUMAN | P02655 | 3 |
| Centromere-associated protein E | A0A087X0P0_HUMAN | A0A087X0P0 | 2 |
| Corticosteroid-binding globulin (Fragment) | G3V4V7_HUMAN | G3V4V7 | 2 |
| Beta-2-microglobulin | B2MG_HUMAN | P61769 | 2 |
| Ig-like domain-containing protein (Fragment) | A0A0J9YY99_HUMAN | A0A0J9YY99 | 2 |
| Protein ITPRID2 | E7EUL7_HUMAN | E7EUL7 | 2 |
| Protein mono-ADP-ribosyltransferase PARP6 (Fragment) | H3BUQ6_HUMAN | H3BUQ6 | 2 |
| Insulin-like growth factor-binding protein complex acid labile subunit | ALS_HUMAN | P35858 | 2 |

**Supplementary Table 6**. Proteins in SNA-enriched Mild COVID-19 Plasma

| Protein Name | UniProt Entry Name | Accession | Average Number of Spectral Matches |
|---|---|---|---|
| Fibrinogen gamma chain | FIBG_HUMAN | P02679 | 25 |
| Complement C3 | CO3_HUMAN | P01024 | 189 |
| Apolipoprotein B-100 | APOB_HUMAN | P04114 | 171 |
| Haptoglobin | HPT_HUMAN | P00738 | 108 |
| Alpha-1-antitrypsin | A1AT_HUMAN | P01009 | 137 |
| Alpha-2-macroglobulin | A2MG_HUMAN | P01023 | 243 |
| Complement factor H | CFAH_HUMAN | P08603 | 104 |
| Haptoglobin-related protein | HPTR_HUMAN | P00739 | 70 |
| Plasminogen | PLMN_HUMAN | P00747 | 39 |
| Hemopexin | HEMO_HUMAN | P02790 | 97 |
| Alpha-1-antichymotrypsin | AACT_HUMAN | P01011 | 35 |
| Kininogen-1 | KNG1_HUMAN | P01042 | 53 |
| Immunoglobulin heavy constant mu | IGHM_HUMAN | P01871 | 60 |
| Inter-alpha-trypsin inhibitor heavy chain H4 | ITIH4_HUMAN | Q14624 | 43 |
| Antithrombin-III | ANT3_HUMAN | P01008 | 49 |
| Apolipoprotein E | APOE_HUMAN | P02649 | 14 |
| Serum amyloid A-1 protein | SAA1_HUMAN | P0DJI8 | 2 |
| Complement factor H-related protein 1 | FHR1_HUMAN | Q03591 | 19 |
| Lipopolysaccharide-binding protein | LBP_HUMAN | P18428 | 11 |
| Hemoglobin subunit alpha | HBA_HUMAN | P69905 | 17 |
| Alpha-2-HS-glycoprotein | FETUA_HUMAN | P02765 | 27 |
| SAA2-SAA4 readthrough | A0A096LPE2_HUMAN | A0A096LPE2 | 8 |
| Inter-alpha-trypsin inhibitor heavy chain H1 | ITIH1_HUMAN | P19827 | 12 |
| Alpha-2-antiplasmin | A2AP_HUMAN | P08697 | 10 |
| Complement factor I | G3XAM2_HUMAN | G3XAM2 | 10 |
| Pregnancy zone protein | PZP_HUMAN | P20742 | 16 |
| Heparin cofactor 2 | HEP2_HUMAN | P05546 | 10 |
| Serum paraoxonase/arylesterase 1 | PON1_HUMAN | P27169 | 5 |
| Afamin | AFAM_HUMAN | P43652 | 23 |
| Plasma kallikrein | KLKB1_HUMAN | P03952 | 7 |
| Apolipoprotein L1 | APOL1_HUMAN | O14791 | 4 |
| Plasma serine protease inhibitor | IPSP_HUMAN | P05154 | 3 |
| Thyroxine-binding globulin | THBG_HUMAN | P05543 | 5 |
| Immunoglobulin lambda-like polypeptide 5 | IGLL5_HUMAN | B9A064 | 5 |
| Apolipoprotein C-II | APOC2_HUMAN | P02655 | 4 |
| Beta-2-microglobulin | B2MG_HUMAN | P61769 | 2 |
| Ig-like domain-containing protein (Fragment) | A0A0J9YY99_HUMAN | A0A0J9YY99 | 5 |
| Protein ITPRID2 | E7EUL7_HUMAN | E7EUL7 | 3 |

**Supplementary Table 7**. Upregulated Proteins in SNA-enriched Severe COVID-19 Plasma

| Protein Name | UniProt Entry Name | Accession | Average Number of Spectral Matches (Severe) | Average Number of Spectral Matches (Mild) |
|---|---|---|---|---|
| Fibrinogen beta chain | FIBB_HUMAN | P02675 | 496 | 115 |
| Fibrinogen alpha chain | FIBA_HUMAN | P02671 | 292 | 91 |
| Fibrinogen gamma chain | FIBG_HUMAN | P02679 | 264 | 25 |
| Haptoglobin | HPT_HUMAN | P00738 | 165 | 108 |
| Fibrinogen gamma chain (Fragment) | C9JPQ9_HUMAN | C9JPQ9 | 136 | 0 |
| Haptoglobin-related protein | HPTR_HUMAN | P00739 | 91 | 70 |
| Plasminogen | PLMN_HUMAN | P00747 | 81 | 39 |
| Alpha-1-antichymotrypsin | AACT_HUMAN | P01011 | 55 | 35 |
| Prothrombin | THRB_HUMAN | P00734 | 51 | 0 |
| Complement component C9 | CO9_HUMAN | P02748 | 29 | 4 |
| Apolipoprotein E | APOE_HUMAN | P02649 | 26 | 14 |
| Vitronectin | VTNC_HUMAN | P04004 | 24 | 9 |
| Complement component C6 | CO6_HUMAN | P13671 | 24 | 2 |
| Serum amyloid A-1 protein | SAA1_HUMAN | P0DJI8 | 24 | 2 |
| Clusterin | CLUS_HUMAN | P10909 | 23 | 12 |
| Complement C4-A | CO4A_HUMAN | P0C0L4 | 22 | 0 |
| Complement C5 | CO5_HUMAN | P01031 | 19 | 3 |
| Complement factor H | A0A0D9SG88_HUMAN | A0A0D9SG88 | 19 | 0 |
| Gelsolin | GELS_HUMAN | P06396 | 18 | 4 |
| Thrombospondin-1 | TSP1_HUMAN | P07996 | 18 | 3 |
| Centrosomal protein of 290 kDa (Fragment) | A0A5K1VW81_HUMAN | A0A5K1VW81 | 16 | 5 |
| Inter-alpha-trypsin inhibitor heavy chain H1 | ITIH1_HUMAN | P19827 | 15 | 12 |
| Complement subcomponent C1r | A0A3B3ISR2_HUMAN | A0A3B3ISR2 | 13 | 0 |
| Complement component 8 subunit beta | F5GY80_HUMAN | F5GY80 | 13 | 1 |
| Serum amyloid P-component | SAMP_HUMAN | P02743 | 11 | 5 |
| Integrin alpha-IIb | ITA2B_HUMAN | P08514 | 10 | 0 |
| Complement component C7 | CO7_HUMAN | P10643 | 10 | 2 |
| Complement component C8 gamma chain | CO8G_HUMAN | P07360 | 9 | 1 |
| Uncharacterized protein C9orf43 | CI043_HUMAN | Q8TAL5 | 9 | 0 |
| Complement component C8 alpha chain | CO8A_HUMAN | P07357 | 9 | 1 |
| Complement factor H-related protein 2 | V9GYE7_HUMAN | V9GYE7 | 9 | 0 |
| Fermitin family homolog 3 | URP2_HUMAN | Q86UX7 | 9 | 0 |
| Angiotensinogen | A0A7P0T9S6_HUMAN | A0A7P0T9S6 | 7 | 0 |
| C-reactive protein | CRP_HUMAN | P02741 | 6 | 0 |
| Carboxypeptidase N catalytic chain | CBPN_HUMAN | P15169 | 6 | 0 |
| Ficolin-3 | FCN3_HUMAN | O75636 | 5 | 1 |
| Pigment epithelium-derived factor | PEDF_HUMAN | P36955 | 5 | 0 |
| Pleckstrin | PLEK_HUMAN | P08567 | 5 | 0 |
| Coagulation factor V | FA5_HUMAN | P12259 | 3 | 0 |
| Potassium voltage-gated channel subfamily H member 1 | A0A0S1TJ81_HUMAN | A0A0S1TJ81 | 3 | 0 |
| Integrin beta (Fragment) | H3BM21_HUMAN | H3BM21 | 3 | 0 |
| Corticosteroid-binding globulin (Fragment) | G3V4V7_HUMAN | G3V4V7 | 2 | 0 |
| Protein mono-ADP-ribosyltransferase PARP6 (Fragment) | H3BUQ6_HUMAN | H3BUQ6 | 2 | 0 |
| Insulin-like growth factor-binding protein complex acid labile subunit | ALS_HUMAN | P35858 | 2 | 0 |
